# Kisspeptin System and Menarcheal Age as Predictors of Primary Female Infertility: A Case-Control Study Among Nigerian Women

**DOI:** 10.1101/2024.07.09.24310162

**Authors:** Izuchukwu Azuka Okafor, Oluseun Olugbenga Saanu, Oladapo Olayemi, Akinyinka O. Omigbodun

## Abstract

**Background:** The Kisspeptin system plays a critical regulatory role in female reproductive functions. However, its role is not yet investigated in primary female infertility (PFI).

**Materials and Methods:** This is a case-control study of consenting primarily infertile (54) and fertile (50) Nigerian females aged between 20 and 44 years who sought consult at University College Hospital, Ibadan. Basic clinical and demographic data were obtained from the participants using a clinical proforma. Five ml of blood were collected by venepuncture for kisspeptin, KISS1, and KISS1R gene expression analysis within the plasma using ELISA and RT-qPCR techniques. The menarcheal age and BMI of the cases and controls were also investigated as possible predictors of PFI.

**Result:** There was no significant change in the relative expression of Kisspeptin, KISS1, and KISS1R genes (p>0.05) in the plasma of the primarily infertile women (PIW) compared to the fertile women (FW). PIW with early menarche showed a significantly lower level of Kisspeptin compared to PIW with normal menarche (p=0.03). Plasma Kisspeptin levels in PIW showed a weak negative correlation (r = −.305; p=0.039) and a good predictive model for KISS1 gene expression (p=0.001; r=0.458). There was a significant difference in KISS1 gene expression in PIW when compared based on their menarcheal age categories (p=0.04).

**Conclusion:** Kisspeptin levels, KISS1, and KISS1R gene expression levels in the blood may not be useful for diagnosing PFI. However, menarcheal age should be investigated as an additional diagnostic indicator for PFI.

## Introduction

Infertility, a complex but common disease, is a worldwide problem affecting 8 - 12% of couples during their reproductive lives ^[1]^. Sub-Saharan Africa has been reported to have one of the highest prevalences of primary (>3%) and secondary (11.6%) infertility in the world ^[2]^. This trend can only be changed if the causes of infertility in the population could be identified, especially as Assisted Reproductive Technology (ART) is relatively inaccessible and very expensive for couples in developing countries ^[3]^. The causes of primary infertility (not being able to be pregnant after at least 1 year of having unprotected sex) have been highlighted in clinical studies over the years ^[4,5]^ and grouped mainly as either male factor, female factor, combined or unknown factors vary across different populations ^[1]^. The International Glossary on Infertility and Fertility Care has defined female infertility as *infertility caused primarily by female factors encompassing: ovulatory disturbances; diminished ovarian reserve; anatomical, endocrine, genetic, functional or immunological abnormalities of the reproductive system; chronic illness; and sexual conditions incompatible with coitus* ^[6]^. It accounts for about 37% of infertility cases among couples ^[7]^.

There is mounting evidence that 10% of infertile females have genetic problems ^[8]^. Several studies have tried to establish the genetic basis of female infertility, but some of these findings have been faulted due to sample size issues, contradictory results, and study peculiarities ^[9–12]^. Despite these findings, genetic causes of female infertility remain unidentified for most patients ^[13]^. Identifying the most promising genetic variants, mutations, or polymorphisms may provide clinically relevant leads with potential for therapeutic intervention, for primarily infertile women (PIW). As additional genes are discovered, and the causes of some infertility disorders become better understood, the management and treatment of female infertility could be improved.

Genome-wide association studies suggest that genetic causes of female infertility could be rare mutations in specific genes and common variations at many sites in the genome, each with a small effect but cumulating in the condition ^[9]^. A critical challenge is the significant number of potential genes that may have an influence on human reproductive function, and which contribute to female infertility. Only a few numbers of genes or genetic disorders are currently substantially linked to infertility. The scenario is fast changing because of the human genome’s completion and advancements in personalized medicine. In fact, 10 to 15 new gene tests are often introduced each year to the list of clinical genetic assays ^[13]^. Thus, a detailed investigation of the genetic basis of female fertility will provide crucial information for the prevention, diagnosis, and treatment of female infertility.

Our understanding of the physiology of the hypothalamic-pituitary-gonadal axis, reproduction, and fertility was fundamentally altered by the discovery of Kisspeptin ^[14–16]^, which acts as a ligand for KISS1R (GPR-54) ^[17,18]^ and produced by the KISS1 gene after a proteolytic process ^[19]^. The KISS1 gene was first identified as a metastasis-suppressing gene before its discovery as a major player in reproductive physiology. ^[17]^

It has been established that Kisspeptin-KISS1R signalling directly affects how GnRH secretion is regulated. Thus, Kisspeptin and KISS1R’s interaction is crucial for controlling the start of puberty and the hypothalamic-pituitary-ovarian axis ^[20,21]^. Inability to reach pubertal development, defective gametogenesis, and a lack of an estrous or menstrual cycle in both mice and humans respectively are some of the reproductive features reportedly caused by mutations in the KISS1R gene ^[22,23]^. Deletions and mutations of KISS1R are associated with severe deficits in the secretion of gonadotropins including luteinizing hormone (LH) and follicle-stimulating hormone (FSH) ^[22,24]^. Similar flaws have also been discovered in KISS1 knockout (KO) mice ^[16,25]^. In humans, Polycystic Ovarian Syndrome (PCOS), a female infertility risk factor, has been associated with low Kisspeptin levels and anomalies of both KISS1 and KISS1R genes ^[26]^. Kallman syndrome and isolated hypogonadotropic hypogonadism have both been linked to KISS1R dysfunction (27,28). Infertility patients may potentially benefit from genetic molecular diagnosis involving the Kisspeptin system if more genetic studies show evidence of its function in human reproduction. This study investigated the expression patterns of KISS1 and KISS1R genes in the plasma of PIF compared to fertile females to determine the relationship between the pattern of expression of these genes and PFI.

## Materials and Methods

### Study design and location

This study is an age-matched case-control study of Kisspeptin, KISS1 and KISS1R gene expression patterns in PIF. This study was carried out in the University College Hospital (UCH) Ibadan, Nigeria as a part of a larger study on the role of Kisspeptin system in primary female infertility amongst Southwestern Nigerian women. The study location was chosen by purposive sampling, while the study participants were recruited by random selection of patients who visited the Gynecology and Family Planning clinic of UCH Ibadan and met the study’s inclusion criteria for both the cases and control groups respectively.

### Sociodemographic, anthropometric and clinical characteristics of study participants

The sociodemographic, anthropometric and clinical characteristics of the study participants were assessed using a clinical proforma. The development, validation and administration of this proforma has been documented in our earlier study together with the associated participants’ characteristics ^[27]^.

### Determination of menarcheal age and BMI of study participants

Menarcheal age and BMI of study participants were also assessed as proportional and continuous variables using a clinical proforma and has been fully described in our previous study ^[27]^. Menarcheal age classification used in this study was according to Glueck et al. ^[28]^: early menarche (≤10 years), normal menarche (11-15 years), and late menarche (≥16 years); while the BMI Classification used was based on the WHO and CDC classification: underweight (<18.5), normal weight (18.5-24.9), overweight (25-29.9), obese (30-34.9), severely obese (35-39.9), morbidly obese (>40). However, this present study established the relationship between the Kisspeptin, KISS1, and KISS1R and how their relationship with menarcheal age and BMI of PIF and fertile females.

### Sample size determination

The sample size for the KISS1 and KISS1R gene expression was determined using the Schlesselman equation for case-control studies and the addition of 10% to cover for attrition ^[29]^. Considering an existing study (30) for sample size determination, the minimum sample size required for this study was 22 subjects each for both case and control groups. G-power software version 3.1 was used to determine the minimum sample size needed to determine significant statistical differences in the plasma Kisspeptin levels of the cases and control. The sample size with an allocation ratio of 1 was determined as two-way statistical testing at the power of 80% and degree of freedom of 0.6 with α probability of error of 0.05. This generated a sample size of 45 each for the cases and controls. However, with the 10% attrition ~ 5, the total sample size required for the study of the plasma Kisspeptin levels is 45 +5 = 50 each for the case and control groups.

### Participant recruitment and selection criteria

The study recruited females diagnosed with primary infertility as cases, while the control subjects were apparently healthy and fertile females. The study participants were between 20 to 44 years old and were Nigerian citizens with no history of foreign phylogeny. For the cases, only females who have never conceived and have regular (2 - 3 days per week) and unprotected intercourse with their partners were recruited. For the controls, only apparently healthy females who have had at least a live birth within the last year (as at the time of recruitment) were recruited.

### Blood sample collection

The WHO phlebotomy protocol was followed in this study ^[31]^. A well-trained phlebotomist was recruited to collect a single 5 ml blood sample by venepuncture from both the cases and controls for KISS1, KISS1R and Kisspeptin analysis. The blood samples collected from each study participant were divided into two portions and put into anticoagulant bottles for ELISA and qPCR analysis, respectively.

### Determination of plasma Kisspeptin levels

The blood plasma Kisspeptin levels were determined using MELSIN Human Kisspeptin ELISA kit (catalog number: EKHU-2171) with an assay range of 0-16 ng/mL, 0.1ng/mL sensitivity, intra-assay CV of <10%, and inter-assay CV of <15%. Patient blood samples were collected in EDTA containers and centrifuged for 30 minutes at 3000g at 2-8°C within 30 minutes. The plasma was retrieved and stored at −20°C until analysis. The stored plasma was thawed, and all sample reagents (20x wash solution - diluted with distilled or deionized water 1:20) were prepared before starting the assay procedure. The standards and samples were added in duplicate to the ELISA plates. The standard and testing sample wells were set, and 50µl and 10µl were added to the standard wells, respectively. 10µl sample diluent was added to the testing sample well, and 100µl of HRP-conjugate reagent. The wells were covered with an adhesive strip, incubated for 60 minutes at 37°C, aspirated, and repeatedly washed five times using a wash solution (400 µl). After the last wash, the remaining wash solution was removed by aspiration or decanting. The plates were inverted and blotted against clean paper towels. Chromogen solution A (50 µl) and Chromogen solution B (50 µl) were added to each well, mixed, and incubated for 15 minutes at 37°C. The plates were protected from light. 50 µl stop solution was added to each well with a gentle tap to the plate for thorough mixing before reading the Optical density (OD) at 450nm using a microtiter plate reader within 15 minutes. The standard curve was used to determine the amount of Kisspeptin in the samples. The standard curve was generated by plotting the average OD (450nm) obtained for each of the standard concentrations on the vertical (Y) axis versus the corresponding concentrations on the horizontal (X) axis. The concentration of the samples was determined by locating the OD value on the Y-axis and extending a horizontal line to the standard curve to find a point of intersection. The standard concentration was 16, 8, 4, 2, 1 and 0 ng/mL.

### KISS1 and KISS1R RNA Isolation

RNA was isolated from the blood using the Jena Bioscience total RNA purification Kit (catalog number: PP-210S). To lyse the blood cells, 500 µl of lysis buffer (2-ME added) was added to 200µl of noncoagulating blood sample in a microcentrifuge tube and vortexed for 10 seconds. For column activation, a spin column was placed into a 2 ml collection tube and added a 100 µl activation buffer for centrifugation at 10000g for 30 seconds. The flowthrough was discarded while the lysate was added a 300 µl isopropanol and vortexed. The mixture was then transferred directly into the spin column and centrifuged at 10000 for 30 seconds before discarding the flowthrough. To wash the primary column, 700 µl of blood washing buffer (ethanol added) was applied to the spin column and centrifuged at 10000g for 30seconds, discarding the flowthrough. This step was repeated with a secondary washing buffer (ethanol added) before another 2 minutes of centrifugation at 10000g to remove residual ethanol. To elute the RNA, the spin column was then placed into a DNase/RNase-free microcentrifuge tube and added 40-50 µl elution buffer to the center of the column membrane before incubation for one minute. Centrifugation was performed at 10000g for one minute to elute the RNA. The remaining genomic DNA was removed from the RNA preparations using the Jena Bioscience DNA Removal kit (Catalogue number: PP-219). The integrity of the RNA was verified by 1% agarose gel electrophoresis. The isolated RNA was stored at −20 or −80°C until further analysis.

### KISS1 and KISS1R gene detection using conventional PCR

As a confirmatory test for the presence of the target DNA, a conventional PCR was performed. According to the manufacturer’s specification, the extracted total RNA was retro-transcribed and amplified using One Taq One-Step RT-PCR kit (catalog number NEB E5315S) by New England Biolabs incorporation. The MJ research Peltier thermal cycler polymerase chain reaction machine used selected primers to target lymphocyte genes. The PCR was performed in a 50 μl volume reaction mixture containing 25 μl volume of one Taq one-step reaction master mix (2x), 2 μl volume of One Taq one-step enzyme mix (2x), 2 μl volume of each gene-specific forward primer (10 μM), 2 μl volume of each gene-specific reverse primer (10 μM), 9 μl volume of nuclease-free water and 10 μl volume of the RNA template was added. Negative control samples for the RT-PCR consisted of a mixture to which all reagents were added except RNA. The PCR was started immediately as follows: Reverse transcriptase at 48°C for 30 seconds, initial denaturation at 94°C for 1 minute, denaturation at 94°C for 15 seconds, annealing at Tm −5°C (the lowest melting temperature of each set of KISS1 gene) for 30 seconds, extension at 68°C for 1 minute, denaturation step for 39 cycles, final extension at 68°C for 5 minutes and final holding at 4°C, forever. The gene nucleotide sequence (5’-3’) for all the primers used are listed in Table 1. The gel was prepared with 1gram of agarose gel powder to 50ml of TBE in a bottle which was melted for 2mins and poured on the plate, while TBE was prepared with 10ml of the stock to 490ml of distilled water. For gel electrophoresis, 1ul of sybr DNA stain was added on a clean foil paper and a 3ul of the template was added and mixed. The mixture was dispensed into the gel-containing wells.

**Table 1:**
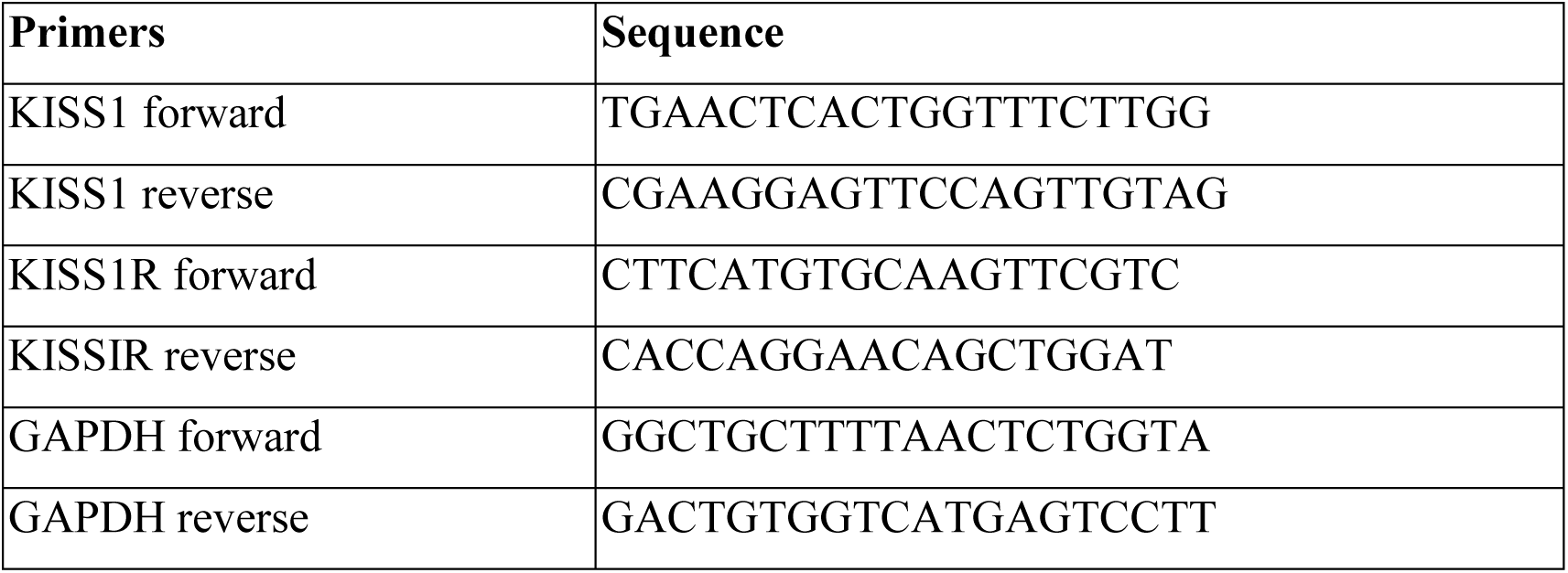
The gene nucleotide sequence for KISS1, KISS1R and GAPDH primers.

### KISS1 and KISS1R cDNA synthesis

The cDNA was synthesized from the isolated RNA using the SCRIPT cDNA synthesis Kit from Jena Bioscience (catalog number: PCR-511.0001). Water, RNA, and primers were mixed before adding other components (Reverse transcriptase, RT buffer, dNTP mix, stock solution, hexamers, oligo-(dT)20 primer, RNase inhibitor, positive control RNA, RNase-free water), which were added to a nuclear-free microtube and mixed by gentle pipetting. To prepare the RNA template/primer mix, RNase-free water, RNA template, and primer were mixed according to the kit’s recommendation and incubated at 65-70°C for 6 mins and placed a room temperature to initiate denaturation and prime annealing. RNase-free water SCRIPT RT, buffer, dNTP mix, DTT stock solution, and RSA inhibitor and sample RT were added to a nuclease-free microtube and mixed. A 10µl was added and mixed before another incubation at 50°C. The quantity and purity of the cDNA were evaluated through spectrophotometry.

### Determination of KISS1 and KISS1R gene Expression

The gene expression levels of KISS1 and KISS1R were determined by quantitative Polymerase Chain Reaction (qPCR) using qPCR SybrMaster (Jena Bioscience PCR-372S, PCR-372L). A master mix was prepared by mixing all the components (qPCR SybrMaster, primer forward, primer reverse, PCR-grade water) as specified in the kit protocol. The master mix was vortexed thoroughly to assure homogeneity and the mix was dispensed into real-time PCR tubes or wells of the PCR plate. Sample/template cDNA (x μl) was added to each reaction vessel containing the master mix, and the tubes were sealed. The microtubes were centrifuged before cycling to remove possible bubbles. Pipet with sterile filter tips were used, and exposure of the master mix to light was minimized. No-template controls were included in all amplifications. The cycle conditions were as follows: Initial denaturation and polymerase activation – x cycles at 95°C for 2mins, as determined optimal; denaturation – 35-45x cycles at 95°C for 15 sec; annealing and elongation - 35-45x cycles at 60-65°C for 1 min. A random expression test was run for a few cases and control samples to estimate significant differences between groups and determine the optimal PCR cycle at which fluorescence gave a better exponential cycle. Sampling and analysis time differences did not exceed 48 hours to ensure the maximum yield of genomic DNA and gene expression.

### Data Analysis

All data were recorded in Microsoft excel, sorted, cleaned, and then transferred to IBM SPSS version 25 for analysis. The group differences in KISS1 and KISS1R gene expression levels and plasma Kisspeptin levels were determined using an independent t-test, while the association between these variables was determined by Spearman correlation and multiple regression analysis. Association between the BMI, menarcheal age and the study participants’ gene expression levels were also determined. All data were screened for normality before analysis using the Shapiro-Wilks’s test. Data were considered significant at p< 0.05.

The similarity in the distribution of mean Kisspeptin levels among the categories of primary infertile women based on BMI and menarche classifications were tested using one-way ANOVA while BMI and menarche subgroup analyses for Kisspeptin, KISS1, and KISS1R gene were analyzed using the Kruskal-Wallis’s test. Post hoc tests were not performed for Kisspeptin levels in PIW because at least one group has fewer than two cases. The group means of Kisspeptin levels and mean fold changes in KISS1 and KISS1R genes were used for the graphical representation of the category distribution analysis instead of the median due to the lack of extreme values and the varying sample sizes of within the sub-groups.

A correspondence analysis with symmetrical normalization was done to determine the relative association between menarche and BMI subtypes in both fertile and primary infertile women. The cross-tabulated frequencies between the variables were used as weight cases for each group analysis. BIPLOT could not be drawn for the corresponding analysis of BMI and menarche in fertile women because there is only one plot dimension because of zero values in some variable categories. For the correspondence analysis of primary infertility whose biplot was drawn, on at least one case of the weight variable, the value was zero. This study did not consider the principal component analysis (PCA) on the Kisspeptin, Kiss1 and Kiss1R gene variables. The exploratory PCA performed with Varimax rotation method showed that observed data did not meet the Kaiser-Meyer-Olkin test for sampling adequacy and showed a low correlation of <0.3 for all components.

### Ethical Considerations

The study protocol was approved by the University of Ibadan/University College Hospital (UI/UCH) Ethics Committee (Assigned number: UI/EC/20/0220; Registration Number: NHREC/05/01/2008a). All the necessary information regarding the study (objectives, requirements of the participants, and duration of the study) were made available to all prospective study participants on an information sheet in English (translated verbally to the native language when requested) to ensure an informed decision to participate in the study. Informed consent was obtained from all the study participants. The study complied with the ethical principles for medical research involving human subjects according to the World Medical Association Declaration of Helsinki.

## Results

### Plasma Kisspeptin, KISS1 gene and KISS1R gene expression levels in fertile and primary infertile women

The mean plasma Kisspeptin levels in both the FW and PIW were not significantly different (p=0.78) (Figure 1). The mean fold relative change in KISS1 and KISS1R gene expressions in both FW and PIW does not vary significantly (p=0.39 and 0.18 respectively) (Figure 1).

**Figure 1:**
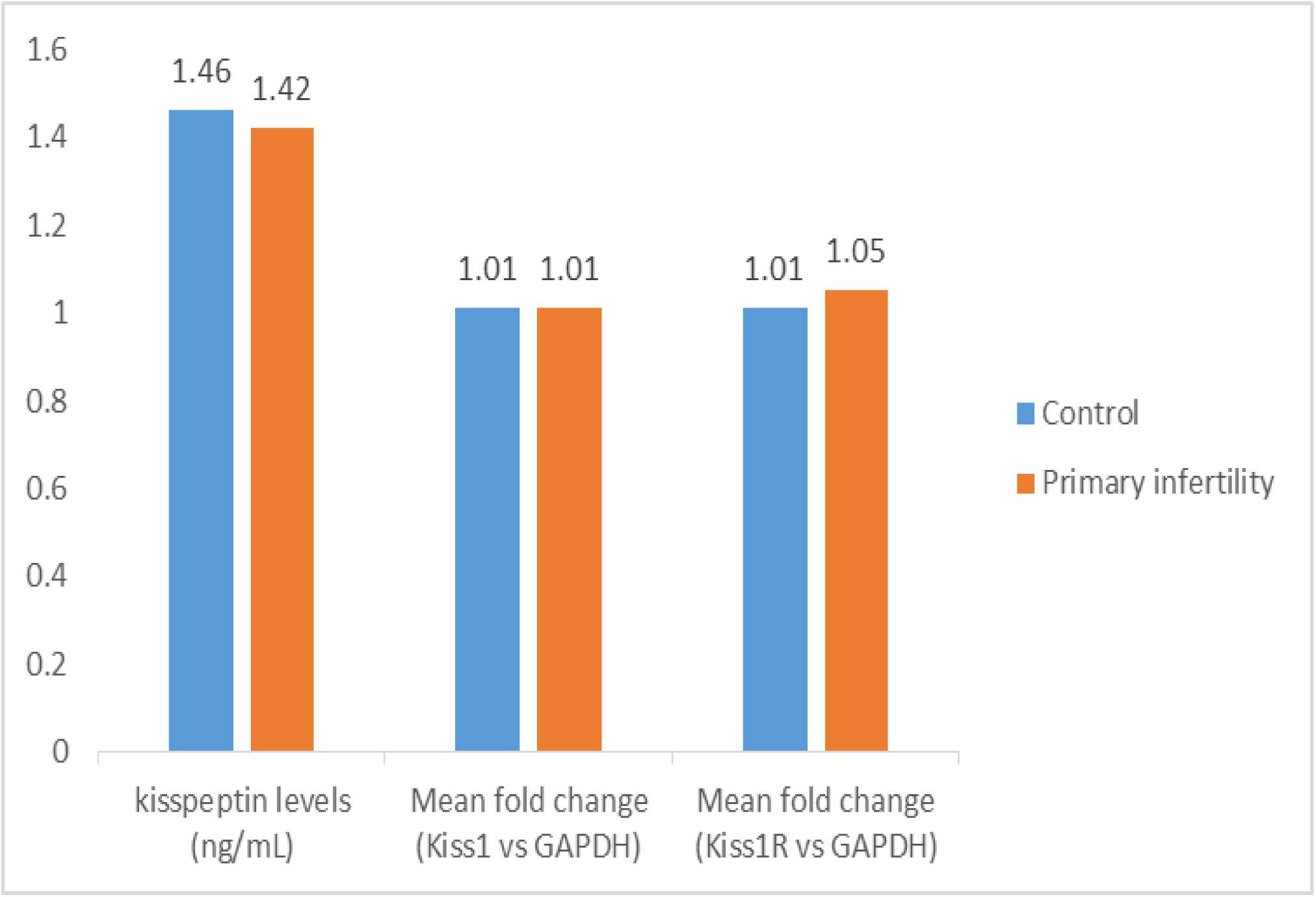
The plasma Kisspeptin levels and mean fold changes of KISS1 and KISS1R relative gene expressions in FW and PIW

### BMI subgroup analysis of Kisspeptin, KISS1 and KISS1R gene expression in fertile and primary infertile women

There was no significant difference in the plasma Kisspeptin levels of FW (p=0.67) and PIW (p=0.56) based on their BMI classifications. The mean fold relative expression of KISS1 gene in FW and PIW based on their BMI classifications were not significantly different (p=0.57 respectively). Also, no substantial change exists in the mean fold relative expression of KISS1R gene in FW (p=0.37) and PIW (p=0.76) based on their BMI classifications (Figure 2–4).

**Figure 2:**
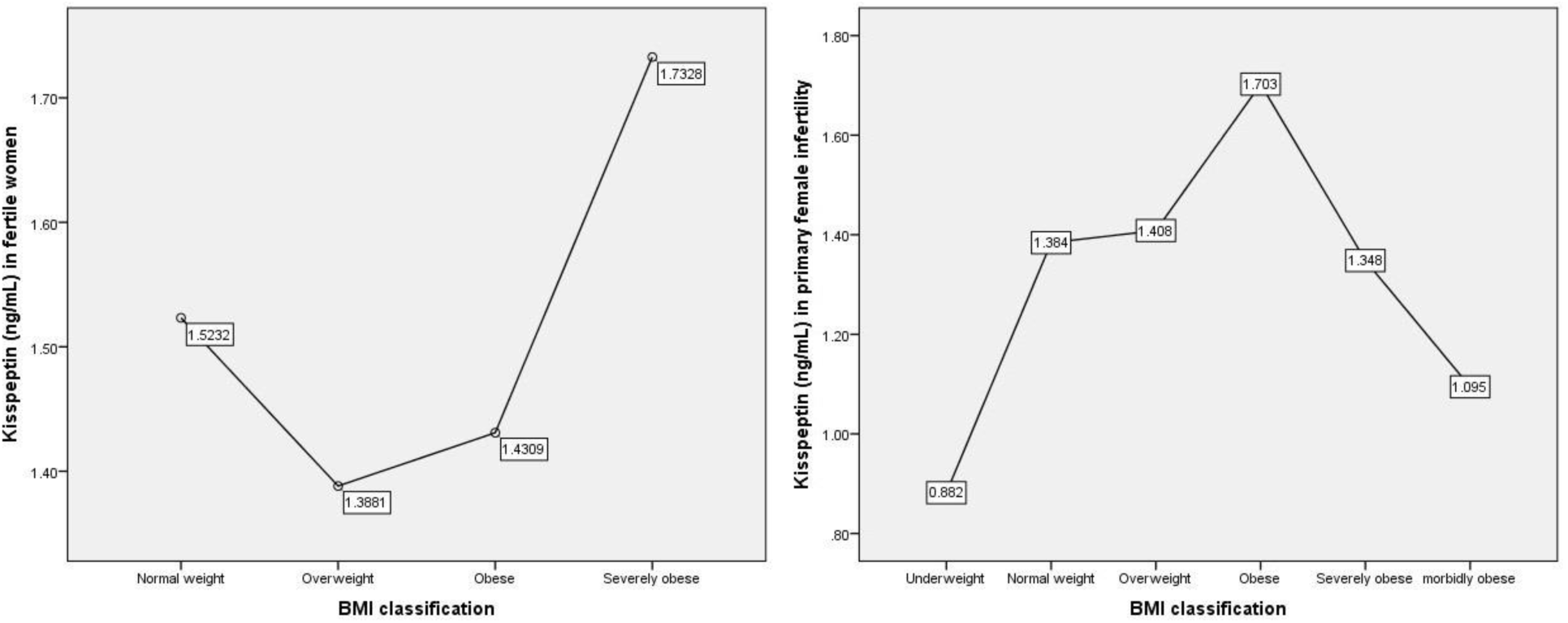
The distribution of mean Kisspeptin levels of FW and PIW according to BMI classification.

**Figure 3:**
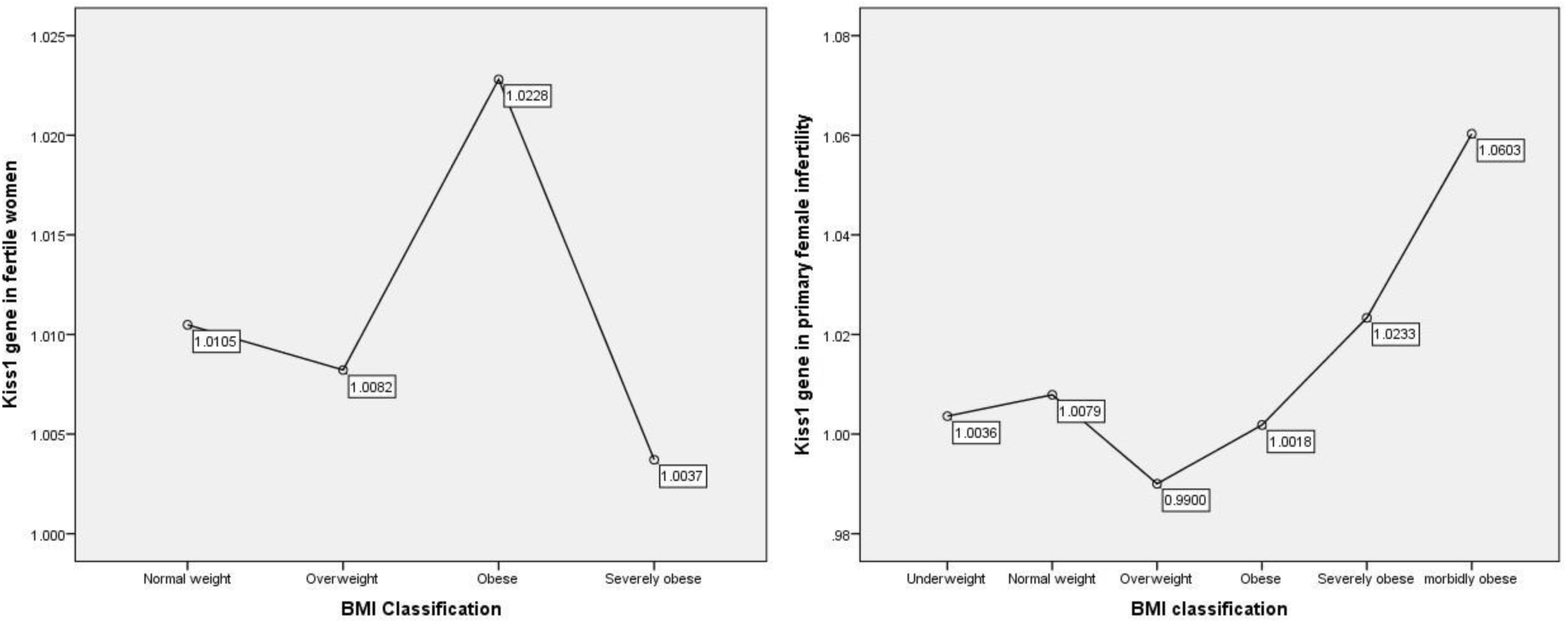
The distribution of KISS1 gene expression in FW and PIW according to BMI classification.

**Figure 4:**
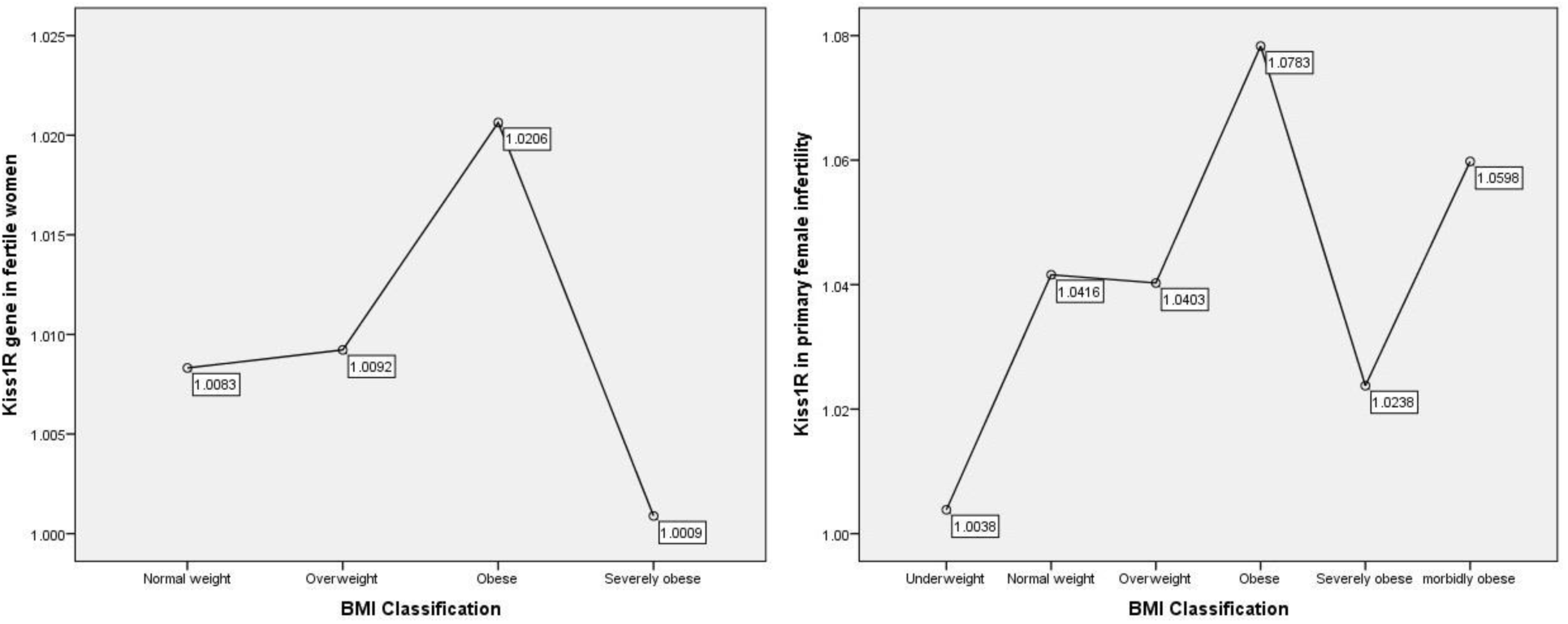
The distribution of KISS1R gene expression in FW and PIW according to BMI classifications.

### Menarche subgroup analysis of Kisspeptin, KISS1 and KISS1R gene expression in FW and PIW

There was no significant difference in the Kisspeptin levels of FW based on their menarche types (p=0.76). However, there was a slightly significant difference in the Kisspeptin levels of PIW based on their menarche categories (p=0.048). PIW with early menarche showed a significantly lower level of Kisspeptin compared to PIW with normal menarche (p=0.03). There was no significant difference in the mean fold relative expression of KISS1 gene in FW based on their menarche types (p=0.48). However, there was a significant difference in the mean fold relative expression of KISS1 gene in PIW according to menarche types (p=0.04). The mean fold relative expression of KISS1R gene in FW (p=0.85) and PIW (p=0.44) based on their menarche types showed no statistically significant difference (Figure 5–7).

**Figure 5:**
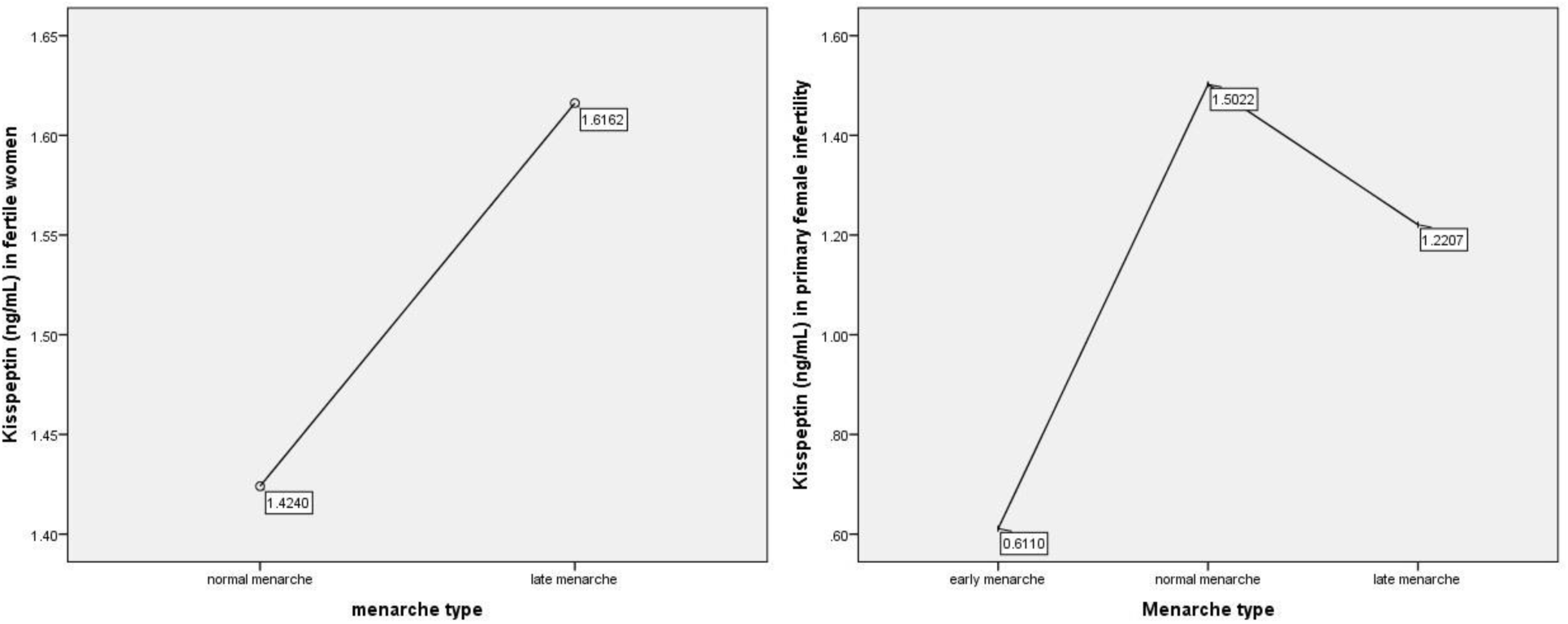
The distribution of mean Kisspeptin levels of FW and PIW according to menarche type.

**Figure 6:**
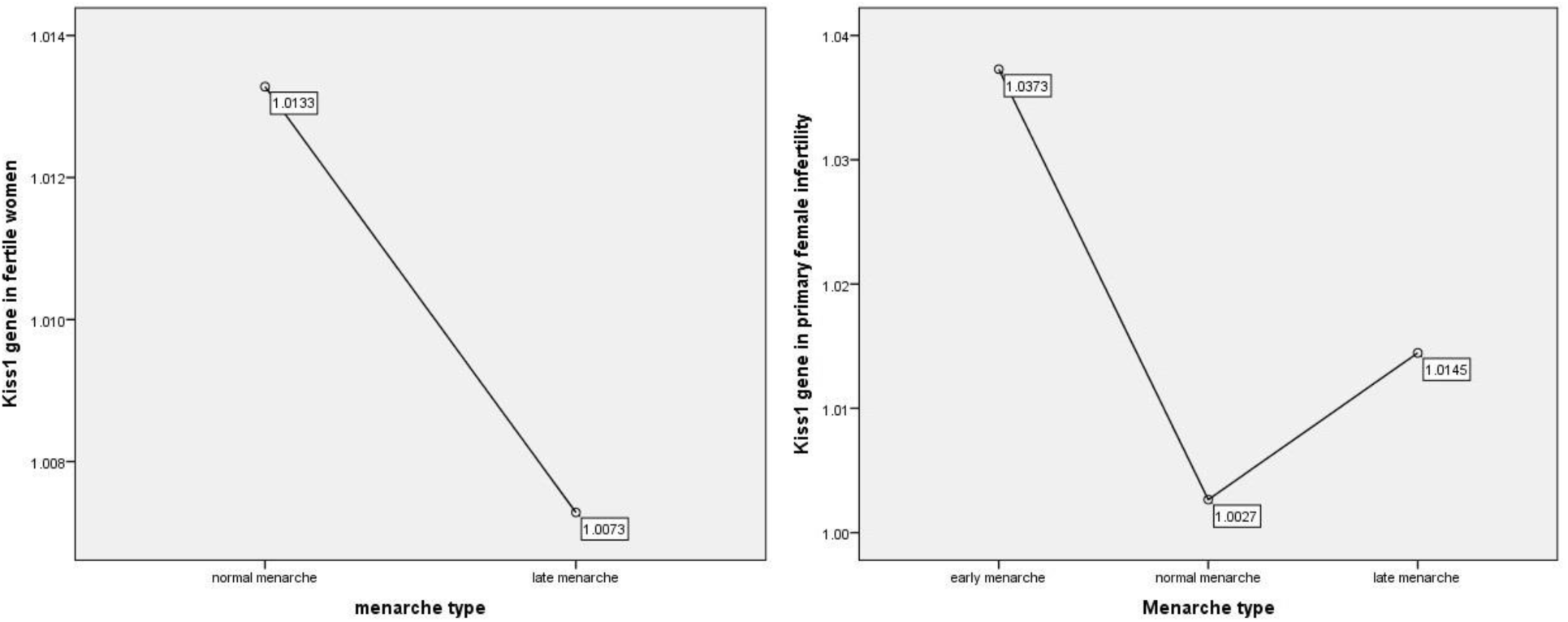
The distribution of KISS1 gene expression in FW and PIW according to menarche type.

**Figure 7:**
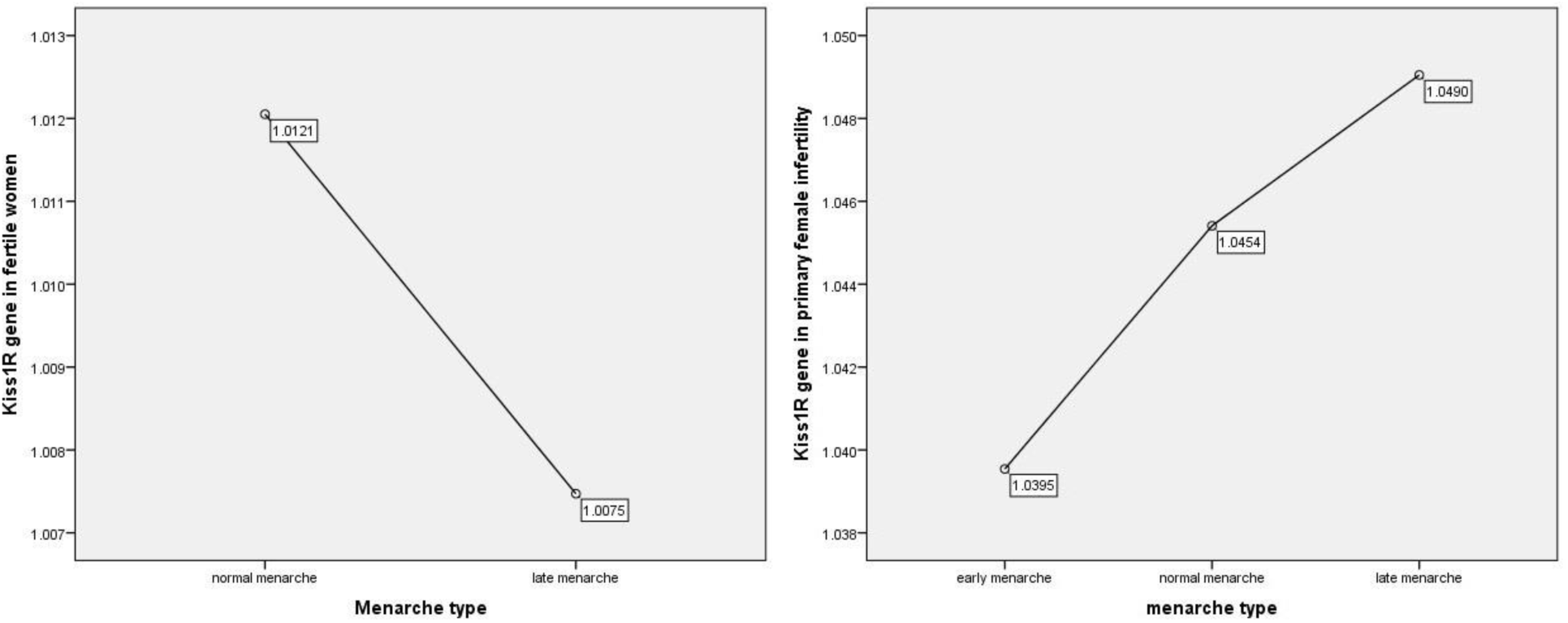
The distribution of KISS1R gene expression in FW and PIW according to menarche type.

### The relationship between BMI, menarche and Kisspeptin system (Kisspeptin, KISS1 and KISS1R genes) in FW and PIW

Table 2 shows the correlation between BMI, menarche and Kisspeptin level, KISS1 and KISS1R genes. There exists no significant correlation (p>0.05) between BMI and Kisspeptin level, KISS1 or KISS1R genes in both FW and PIW. Menarche shows a similar correlation trend (p>0.05) in both FW and PIW, except in fertile women where it shows a slightly significant moderate positive correlation with Kisspeptin levels (r=0.38, p=0.04).

**Table 2:** Correlation between BMI, menarche and Kisspeptin levels, KISS1 and KISS1R genes.

| Parameters |  | Kisspeptin<br>(ng/mL) |  | KISS1 gene |  | KISS1R gene |  |
| --- | --- | --- | --- | --- | --- | --- | --- |
|  |  | FW | PIW | FW | PIW | FW | PIW |
| **BMI**<br>**(m/kg<sup>2</sup>)** | Pearson<br>Correlation | -.044 | .062 | .171 | .136 | .039 | -.022 |
|  | Sig. (2-tailed) | .819 | .681 | .327 | .346 | .822 | .880 |
|  | N | 30 | 46 | 35 | 50 | 35 | 48 |
| **Menarche (years)** | Pearson Correlation | .379 | -.030 | -.122 | .173 | -.072 | -.116 |
|  | Sig. (2-tailed) | **.039** | .845 | .485 | .229 | .683 | .430 |
|  | N | 30 | 46 | 35 | 50 | 35 | 48 |

N = sample size; BMI = body mass index; FW = fertile women; PIW = Primarily infertile women

### Prediction analysis for Kisspeptin levels, KISS1 and KISS1R gene expression with BMI and menarche using multiple regression analysis

The regression prediction of Kisspeptin levels, KISS1 and KISS1 gene expressions with BMI or menarche showed no significant model for both FW and PIW (p>0.05) (Table 3).

**Table 3:**
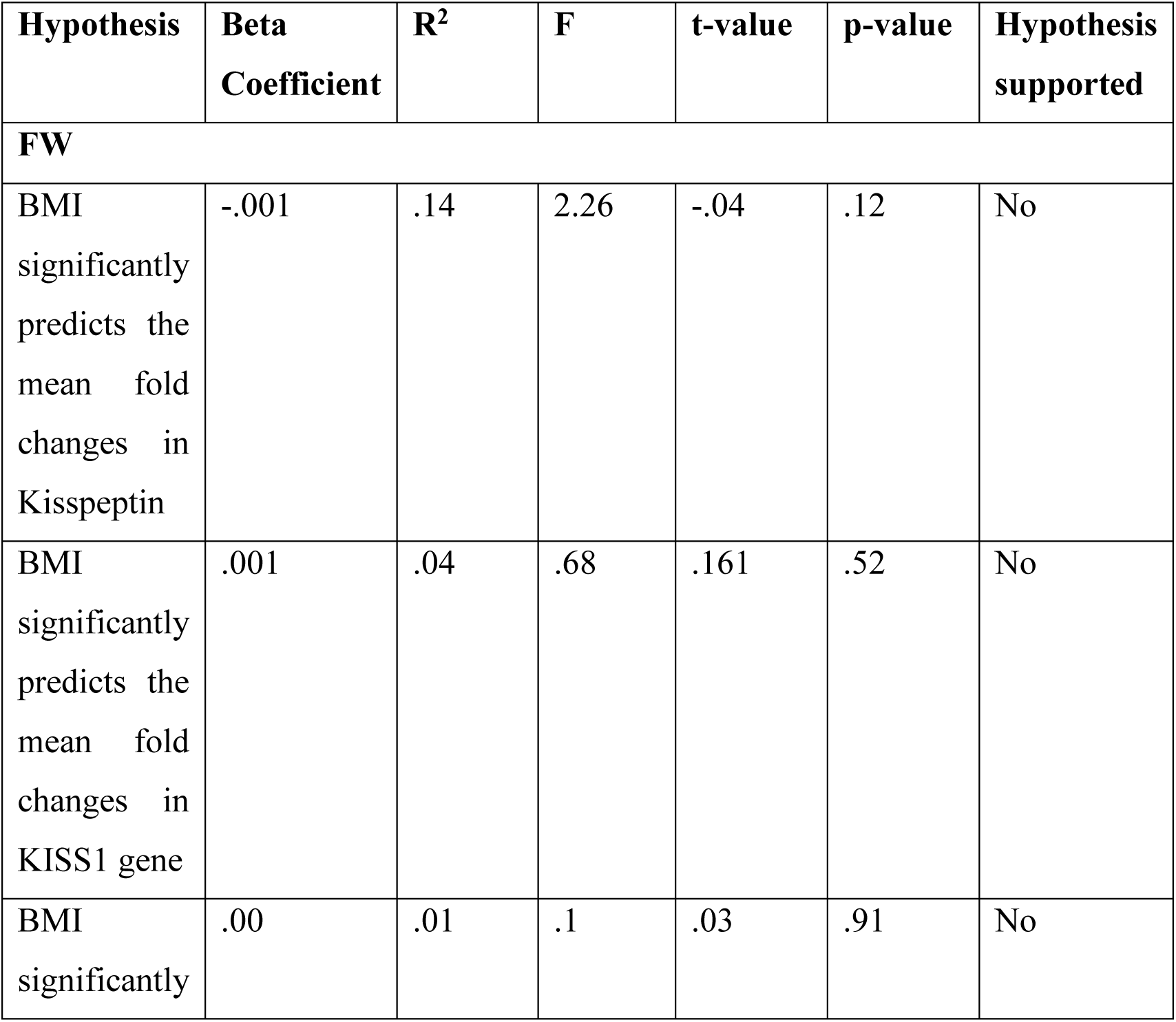
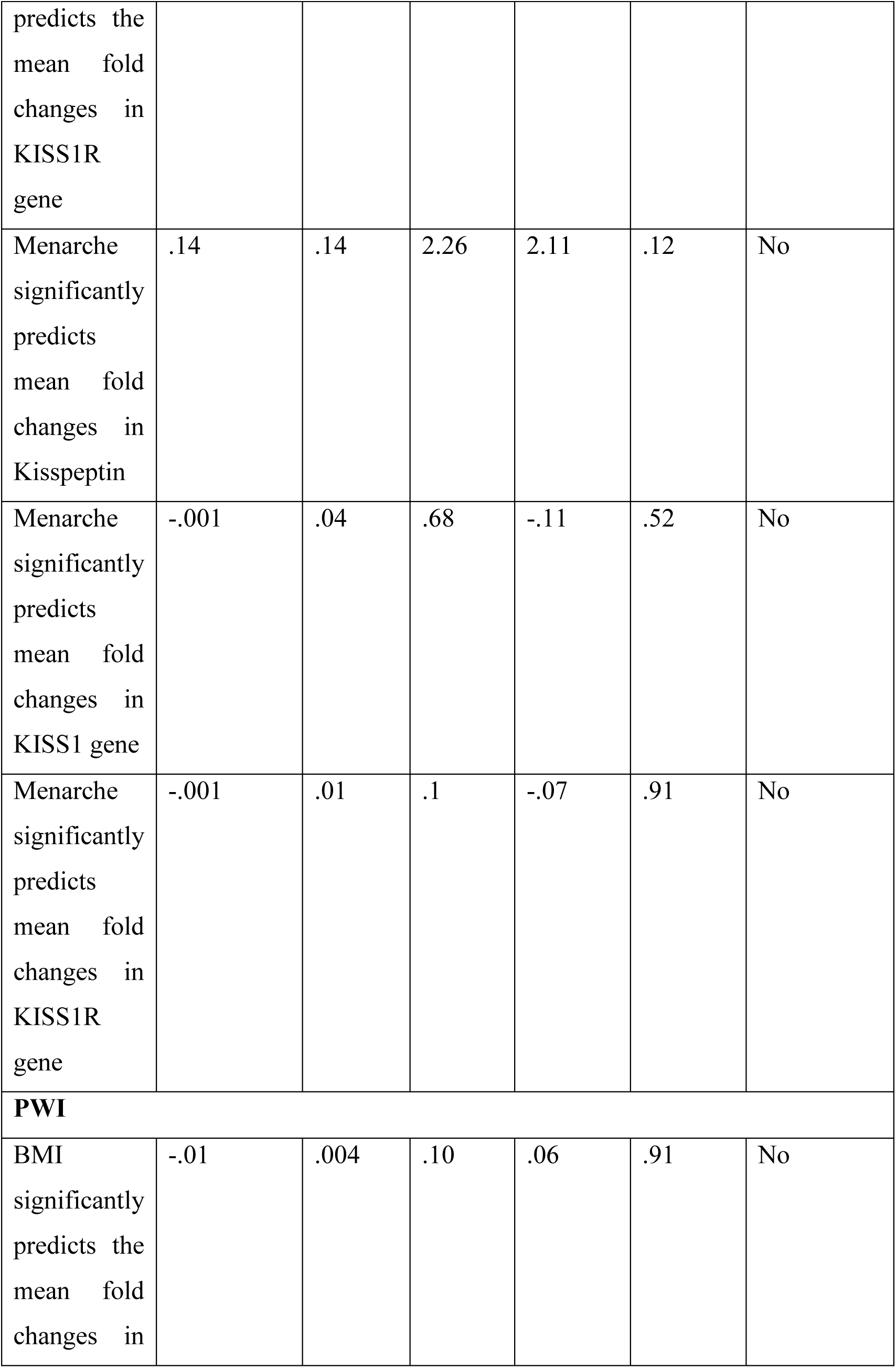
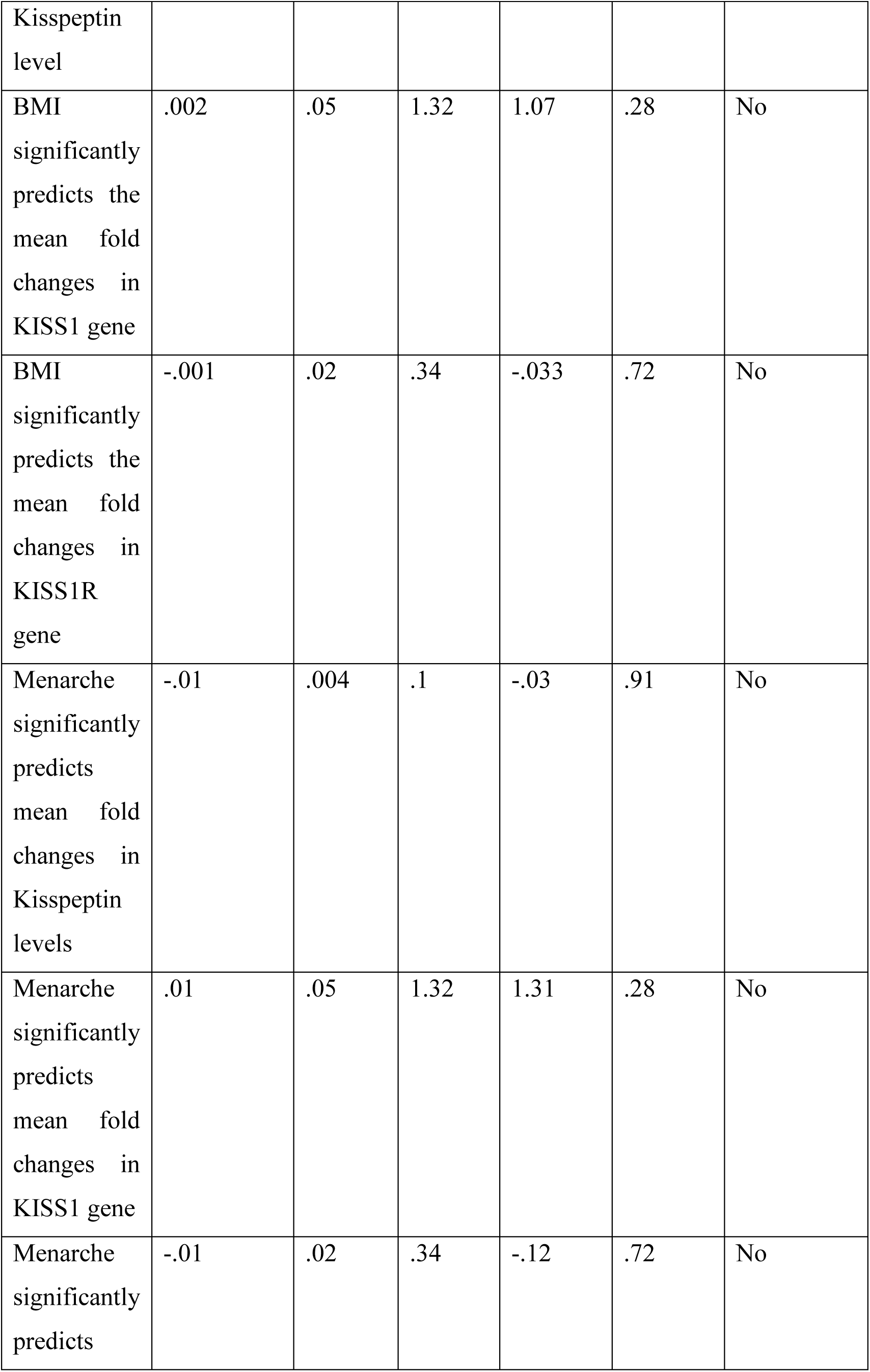
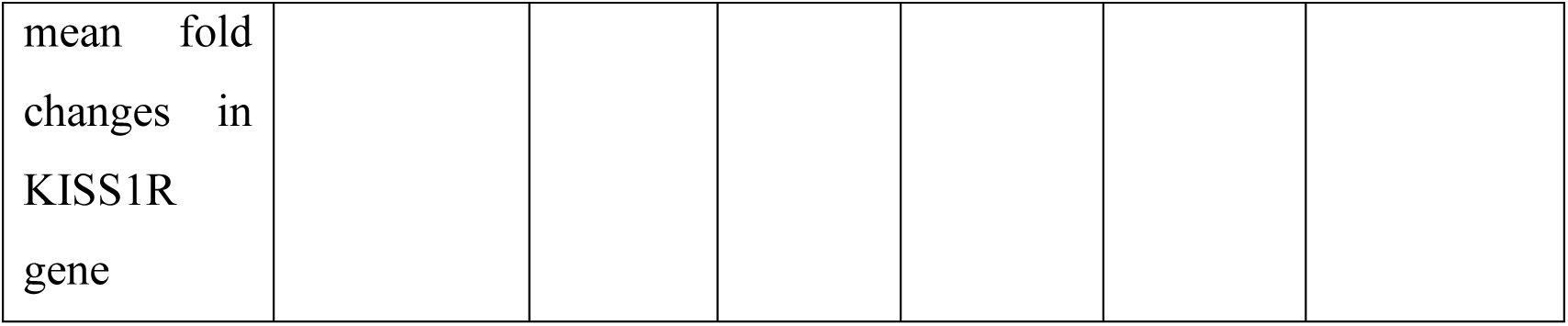
Prediction of Kisspeptin, KISS1 and KISS1R gene expression in FW and PIW using BMI and menarche.

### Correspondence analysis showing the relative association between menarche and BMI subtypes in PIW

The two-dimensional correspondence analysis revealed no relationship between any of the menarcheal categories and BMI subtypes in PIW (Figure 8).

**Figure 8:**
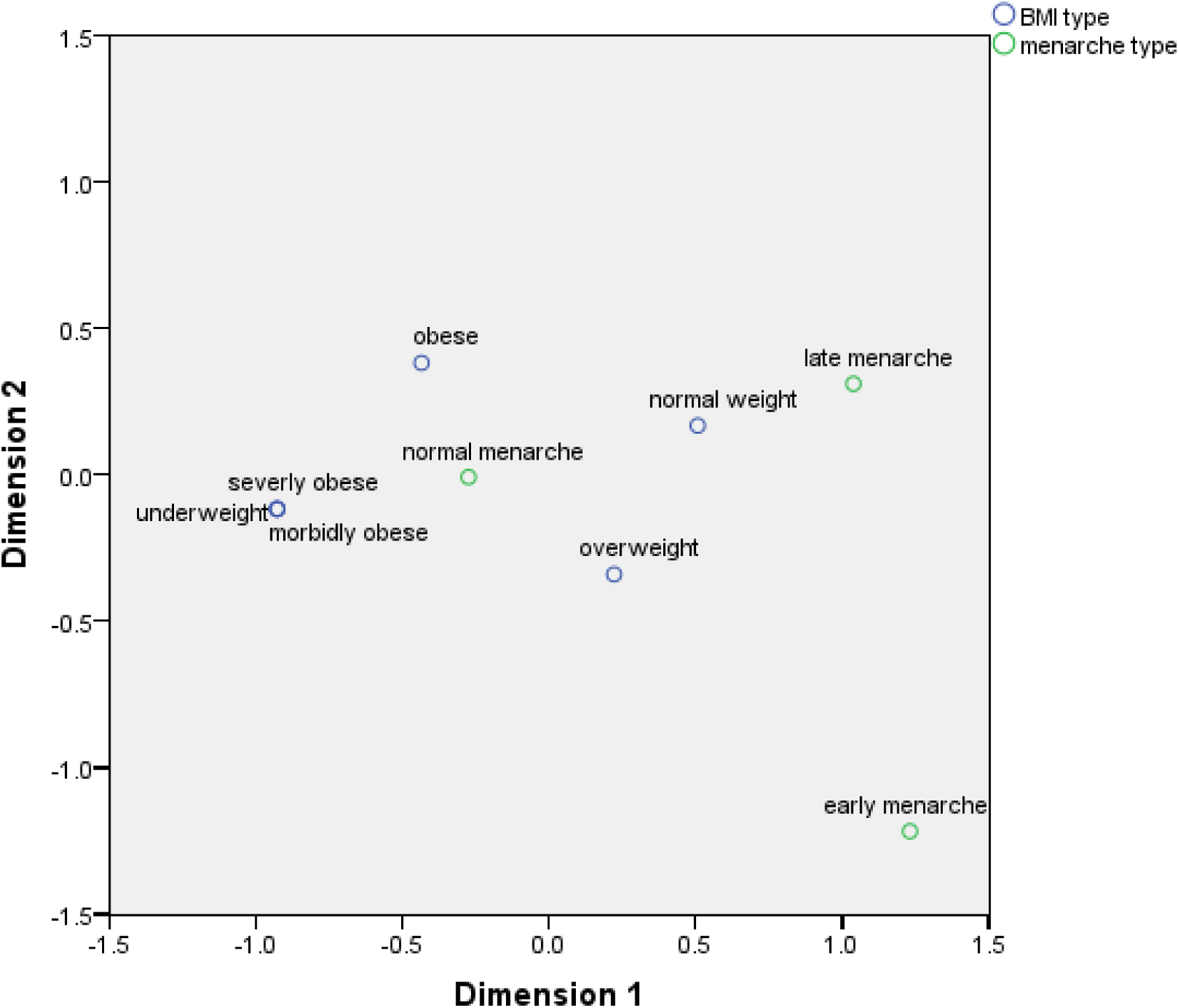
A two-dimension correspondence analysis showing the relative association between menarche and BMI subtypes in PIW

### The relationship between Kisspeptin levels, KISS1 and KISS1R gene expressions in FW and PIW

There was no relationship between Kisspeptin levels and KISS1R in both PIW (r= .05, p = .78) and FW (r=0.09, p= 0.57) (Table 4 and 5). However, there was a weak negative correlation between the Kisspeptin levels and KISS1 gene expression in PIW (r=-.031, p=.04). The KISS1 gene expression in FW showed a significant moderate positive correlation with KISS1R (r=.52, p<0.01) while it showed no significant relationship with KISS1R gene expression in PIW (r=-.09, p=.54).

**Table 4.**
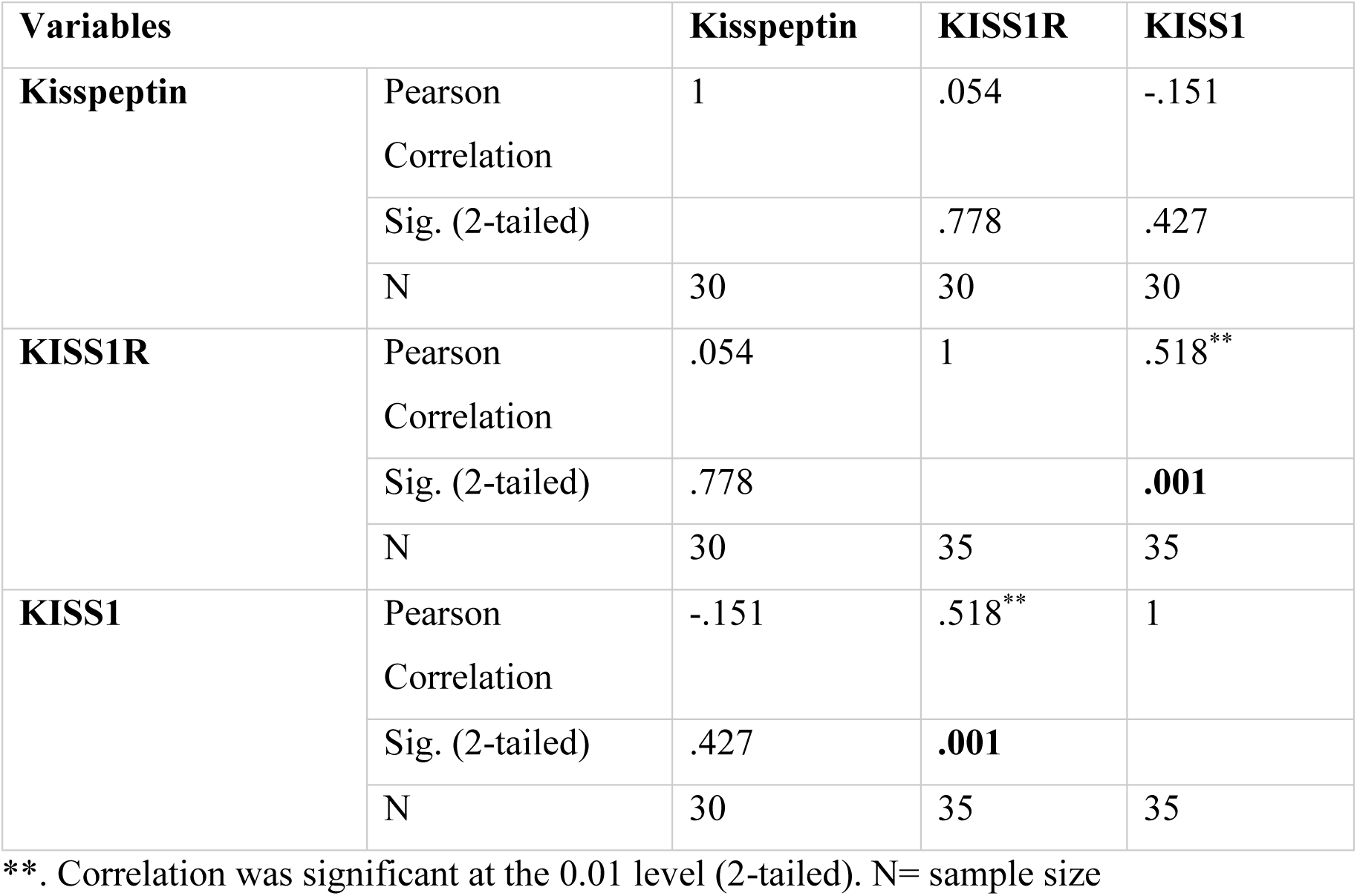
Correlations between Kisspeptin levels, KISS1 and KISS1R gene expression levels in FW.

| Variables |  | Kisspeptin | KISS1R | KISS1 |
| --- | --- | --- | --- | --- |
| **Kisspeptin** | Pearson Correlation | 1 | .054 | -.151 |
|  | Sig. (2-tailed) |  | .778 | .427 |
|  | N | 30 | 30 | 30 |
| **KISS1R** | Pearson Correlation | .054 | 1 | .518** |
|  | Sig. (2-tailed) | .778 |  | .001 |
|  | N | 30 | 35 | 35 |
| **KISS1** | Pearson Correlation | -.151 | .518** | 1 |
|  | Sig. (2-tailed) | .427 | .001 |  |
|  | N | 30 | 35 | 35 |

\*\*. Correlation was significant at the 0.01 level (2-tailed). N= sample size

**Table 5:** Correlations between Kisspeptin levels, KISS1 and KISS1R gene expression levels in PIW.

| Variables |  | Kisspeptin | KISS1R | KISS1 |
| --- | --- | --- | --- | --- |
| **Kisspeptin** | Pearson Correlation | 1 | .089 | -.305* |
|  | Sig. (2-tailed) |  | .565 | **.039** |
|  | N | 46 | 44 | 46 |
| **KISS1R** | Pearson Correlation | .089 | 1 | -.090 |
|  | Sig. (2-tailed) | .565 |  | .544 |
|  | N | 44 | 48 | 48 |
| **KISS1** | Pearson Correlation | -.305* | -.090 | 1 |
|  | Sig. (2-tailed) | **.039** | .544 |  |
|  | N | 46 | 48 | 50 |

\*. Correlation is significant at the 0.05 level (2-tailed). N= sample size

### Multiple regression and prediction analysis of KISS1 gene expression using Kisspeptin and KISS1R gene expression

The prediction of mean fold changes in KISS1 gene expression in FW using mean plasma Kisspeptin levels and mean fold change in KISS1R gene showed a significant model [F (2,27) =5.45; p=.01] with only 29% of the variations in KISS1 gene expression (R^2^ = .29) being explained by changes in Kisspeptin levels and KISS1R gene expression. Mean fold changes in KISS1R was a significant predictor of mean fold changes in KISS1 gene expression in FW (p=.004) with a unit change in KISS1R expression causing 0.58-unit increase in KISS1 gene expression (B=0.58) (Table 6).

**Table 6:**
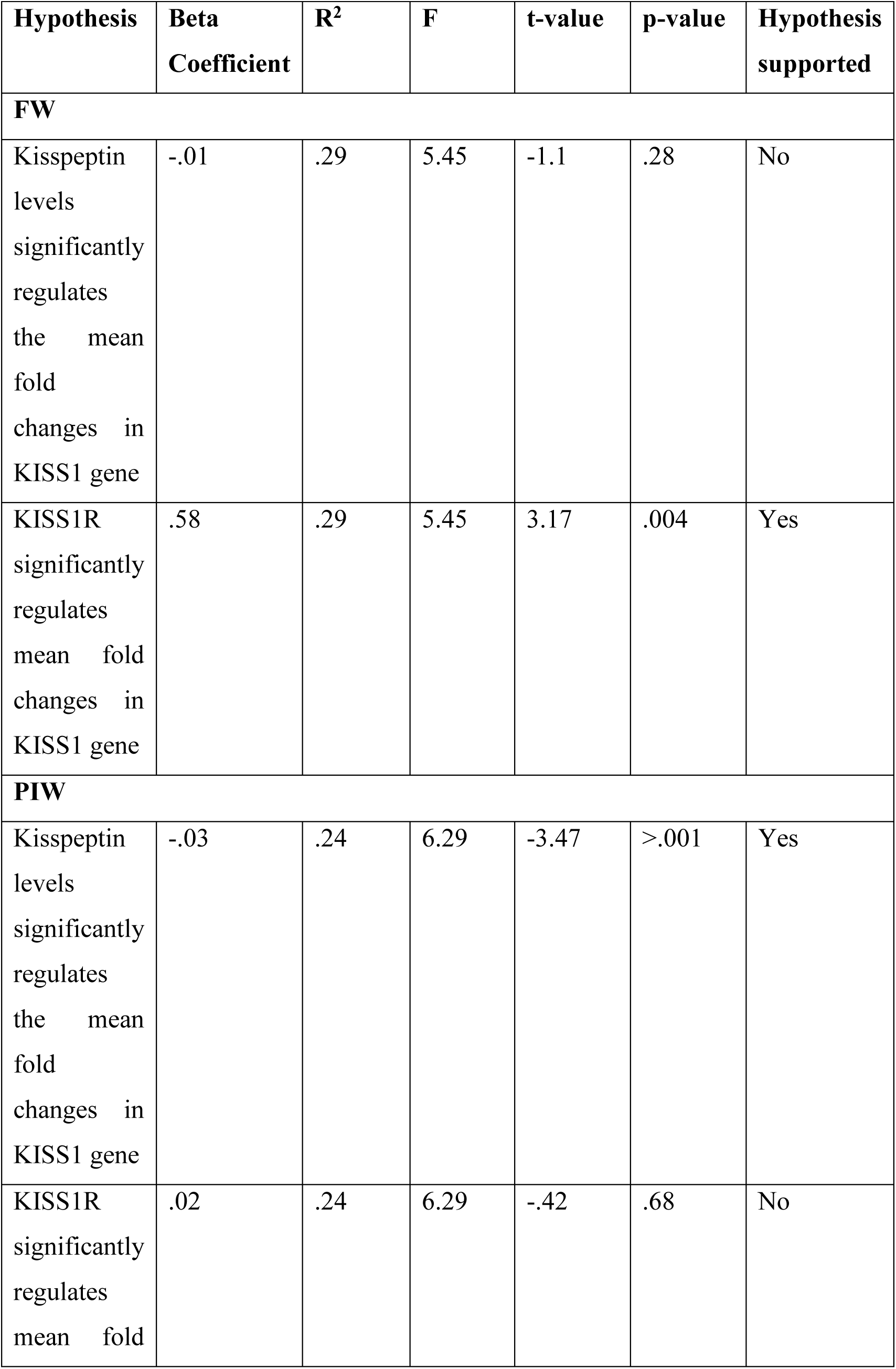
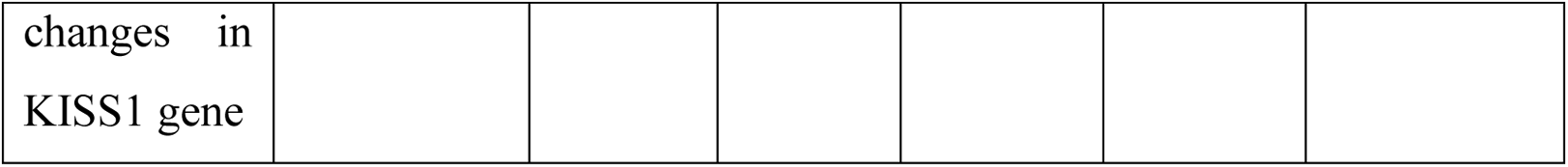
Prediction of KISS1 gene expression using mean Kisspeptin levels and mean fold changes in KISS1R gene expression.

The prediction of mean fold changes in KISS1 gene in PIW using mean plasma Kisspeptin levels and mean fold changes in KISS1R gene showed a significant model [F (2,41) =6.29; p=.004] with 24% of the variations in KISS1 gene expression (R^2^ = .24) being explained by changes in Kisspeptin levels and KISS1R gene expression. Mean Kisspeptin levels was a significant predictor of the mean fold changes in KISS1 gene of primary infertile women (p>.001) with a unit change in Kisspeptin levels causing 0.03-unit decrease in KISS1 gene expression (B=-0.03) (Table 6).

## Discussion

This study observed the changes in the Kisspeptin system (Kisspeptin, KISS1 gene, and KISS1R gene) in the plasma of PIW compared to FW (Figure 1). The Kisspeptin levels in the plasma of PIW compared to the FW were comparable and did not differ significantly. This is the first known study reporting Kisspeptin levels in PIW. However, some studies reported the level of Kisspeptin in PCOS, one of the clinical diagnoses of PFI ^[32–36]^. Many of these studies fail to classify their patients according to the type of infertility – one could assume that the population is mixed - making it difficult to compare with this present study. Howbeit, it may be beneficial to use the data on PCOS - being an important factor in PFI – to contextualize the findings of this present study. It is important to note that available studies investigated the Kisspeptin levels of PCOS women in both serum and plasma. The fact that the degradation of Kisspeptin in the serum varies compared to plasma may have also affected the outcome of these studies ^[37]^.

Recent research indicates that women with PCOS have increased Kisspeptin levels and a higher LH/KP ratio ^[33,36]^. A systematic review reported increased Kisspeptin levels in the serum or plasma of eight out of twelve studies reviewed ^[38]^, while a recent meta-analysis of nine studies concluded that there was a statistical difference in the serum Kisspeptin level between PCOS and non-PCOS women ^[39]^. As much as the diagnosis of PFI may not be heterogeneous, the diagnosis of the different causes may also affect the result seen in this present study. The determination of the Kisspeptin level in women with a known cause of PFI may be more insightful than a generalization of PIW. Though the different clinical diagnoses of PFI of participants included in this present study have been earlier reported ^[27]^, the sample sizes of each subgroup were small to allow a scientifically valid comparison of Kisspeptin levels between the FW and PIW.

KISS1 and KISS1R gene expression in the blood equally showed no statistical difference amongst FW and PIW. This finding corroborates the reported Kisspeptin levels in plasma because of the production relationship between the Kisspeptin and KISS1 gene. KISS1 and KISS1R genes have been mainly investigated in other specimens (cumulus cells, decidua cells, trophoblasts, granulosa cells) other than the blood ^[40–46]^. Despite the low detection of KISS1/KISS1R in the blood ^[47]^, this study investigated the KISS1/KISS1R gene expression in the blood, with validated procedures to ensure maximal detection. This is the first study describing KISS1 or KISS1R gene expression in PIW. To be considered a potentially safe and less invasive diagnostic marker for PFI, a blood investigation of KISS1 or KISS1R gene expression should be more appropriate and desired.

Studies which utilized other tissues for investigating KISS1 and KISS1R gene expression showed interesting results. In patients with endometriosis, KISS1 gene expression was not found in any endometrial sample collected ^[48]^. Also, the level of KISS1 expression was statistically substantially greater in endometriosis lesions than in eutopic glandular endometrium, suggesting the role of the KISS1 gene in the etiology of endometriosis ^[48]^ which was earlier reported to be part of the clinical diagnosis of primary infertility in the present study cohort ^[27]^. Similarly, Blasco and colleagues ^[40]^ discovered that endometriosis patients’ cumulus cells had more KISS1R mRNA than healthy oocyte donors’ cumulus cells. As a result, the researchers concluded that elevated KISS1R gene expression might be one of the mechanisms in the etiology of endometriosis and associated infertility. Other studies on placental and decidual tissues showed that women who experienced spontaneous abortion in the first trimester significantly reduced KISS1 gene expression compared to women who underwent elective termination ^[41]^. Kisspeptin and KISS1R expressions in syncytiotrophoblasts and cytotrophoblasts were decreased in women with recurrent spontaneous abortion compared to women with voluntary abortion. While Kisspeptin levels reduced significantly in decidua of these women, there were no significant changes in KISS1R gene expression when women with recurrent spontaneous abortion were compared to women with voluntary abortion ^[43,46]^. In this present study, women who had any records of abortion were excluded as they do not meet the definition of PFI. However, the above findings by previous studies on abortion and Kisspeptin system changes are useful because abortifacient factors are implicated in PFI ^[49,50]^.

In this present study, there exists no difference in the distribution of Kisspeptin levels, KISS1 and KISS1R gene expression following the subgroup analysis of FW and PIW based on their BMI classification (Figures 2-4). As earlier published ^[27,51]^, more than half of the study participants have an abnormal BMI classification, with their BMI average being in the overweight category. There was no relationship between the BMI of both FW and PIW and any of Kisspeptin, KISS1, and KISS1R gene expression levels (Table 2 and 3). This finding is comparable with a study of Saudi females with normal and abnormal weight, which found no correlation between BMI and serum Kisspeptin level. They also reported that Kisspeptin levels in the overweight/obese group did not differ significantly with women with normal weight. However, the subjects were not reported to have any form of infertility ^[52]^. In another study involving females with PCOS, there were significantly decreased Kisspeptin levels in underweight, overweight, and obese women compared to women with normal weight groups. More importantly, Kisspeptin levels decreased with increase in BMI ^[53]^. A recent review created a network of functional association between Kisspeptin and obesity-related genes and identified gonadotropin-releasing hormone 1 (GNRH1) as one of the twelve signalosome hubs which will provide novel insight into the body’s energy homeostasis. An earlier study on mice showed evidence that Kisspeptin signaling influences body weight, energy expenditure, and glucose homeostasis in a sexually dimorphic and partially sex steroid-independent fashion; therefore, changes in Kisspeptin signaling might contribute, to some aspects of human obesity ^[54]^. Hence, future studies should focus on determining where the metabolic effects of Kisspeptin signaling occur in humans and if similar phenotypes exist in humans who do have decreased expression or non-functional Kisspeptin, KISS1 and KISS1R genes ^[55]^.

The Kisspeptin system plays a role in puberty development, which menarche represents, through its direct regulation of GnRH ^[56]^. Precocious puberty has been associated to increased Kisspeptin secretion ^[57–60]^ whereas this present study shows a reduced Kisspeptin level for PIW with early menarche compared to PIW with normal menarche (Figure 5). In contrast, PIW with early menarche had a significantly increased mean fold KISS1 gene expression than PIW with normal menarche (Figure 6). This finding relates to the slightly significant moderate positive correlation between menarcheal age of PIW and their plasma Kisspeptin levels (Table 2). An abnormal puberty development could lead to delayed or early menarche, which could result from Kisspeptin dysregulation ^[61–65]^. Combined, it could be hypothesized that the early onset of puberty induces some form of subfertility in the future, associated with reduced Kisspeptin levels which may depend on some heritable genetic factors ^[66]^.

The findings of this present study add to the justification for Kisspeptin supplementation for GnRH stimulation and suggest that supplementation may be needed by PIW who present with a history of early menarche. Kisspeptin supplementation has been used in clinical trials as KP-10 and KP-54 isoforms in females with hypothalamic amenorrhoea ^[67–69]^, PCOS ^[35]^, or infertile women requiring IVF, with a need to prevent OHSS or stimulate ovulation ^[70–73]^. The trials mentioned above have shown no noticeable side effects across various doses, times, and routes of administration. Kisspeptin can stimulate effective GnRH release and has been optimally functional when administered in the preovulatory period ^[67]^ and with reduced sex steroid ^[68]^. The clinical outcomes and the role of Kisspeptin supplementation in human reproduction have been critically reviewed in the literature highlighting areas for future research and clinical policy considerations ^[74,75]^. There are still no supplementation protocols for clinical settings to date despite the success of Kisspeptin supplementation trials in women.

This present study showed no significant prediction of plasma Kisspeptin level, KISS1 and KISS1R gene expression with BMI or menarcheal age of FW or PIW (Table 3). It is important to note that no relative association exists between the categorized BMI and menarcheal age subtypes in both FW and PIW following a correspondence analysis (Figure 8), which agrees with the earlier finding on the uncategorized BMI and menarcheal age ^[27,51]^.

There exist some linear relationships between the components of the Kisspeptin system based on the findings from this present study. There was a significant weak negative correlation between the KISS1 gene expression and Kisspeptin levels in PIW (r=-.031, p=.04). An inverse relationship was observed between the KISS1 gene and its peptide, Kisspeptin, describing a slight chance for the two variables to move in different directions in PFI. The negative correlation, albeit weak, seen between KISS1 and Kisspeptin could explain the possibilities of other internal or external influences on its expression in the human blood. There is lack of data on the control of the Kisspeptin system in human blood. However, there exists well-documented evidence in animals implicating aging and sex steroids (testosterone, dihydrotestosterone, and estrogen) as major players, with slight variation across species and sites of detection ^[76–81]^. Evidence from rats and mice shows that the circulating levels of estrogen determine the expression level of the Kisspeptin and KISS1 gene ^[76,77]^. This relays the negative and positive feedback control of estrogen on GnRH. There is usually an atypical production of estrogen during premature ovarian failure or premature ovarian insufficiency, which could affect the expression of the KISS1 gene leading to the antagonistic relation between the gene and its encoded protein, Kisspeptin. In this present study, the KISS1 gene expression in FW showed a significant moderate positive correlation with KISS1R gene expression (r=.52, p<0.01), adjudging a similar trend in the changes across the two variables. There may be a potentially identical regulatory effect on KISS1 and KISS1R. In other animals, estrogen and testosterone have no control on the expression of KISS1R compared to their significant impact on the KISS1 gene ^[82,83]^. Hence, studying these regulations in humans is crucial to understanding the true relationship between the Kisspeptin system genes.

This present study utilized a linear multinomial regression model to predict the mean fold change of mRNA levels of the KISS1 gene using Kisspeptin and KISS1R mean fold change. The regression model used was significant in both FW and PIW. While 24% of changes in KISS1 gene expression in PIW result from changes in Kisspeptin and KISS1R, 29% of variations in KISS1 gene expression in FW result from changes in Kisspeptin and KISS1R gene expression levels. The mean fold changes in KISS1R are a significant predictor of mean fold changes in the KISS1 gene of FW (p=.004), with a unit change in KISS1R expression causing a 0.58-unit increase in KISS1 gene expression (B=0.58). However, in PIW, the mean Kisspeptin levels are a significant predictor of the mean fold changes in the KISS1 gene of PIW (p>.001), with a unit change in Kisspeptin levels causing a 0.03unit decrease in KISS1 gene expression (B=-0.03).

One of the significant limiting features of gene expression studies is the propensity for small sample size, even when the sample size was appropriately determined. The evidence submitted in this study relies on a small sample size, especially for the subgroup analysis, where it became impossible to compare some group with another due to zero value. For example, the correspondence analysis biplot could not be drawn for the BMI and menarcheal age association in FW because of zero values in some variable categories. This was the case for many published studies on Kisspeptin system gene expression in different tissues ^[41,43,46,72,84]^. However, this current study is one of the few studies ^[43,85,86]^ that reported the expression of KISS1/KISS1R in the blood or blood cells. Other KISS1/KISS1R expression studies ^[41,43,44,46]^ have focused on other tissues due to the low level of expression of these genes in the blood. This study did not assess the karyotypes of all patients recruited to avoid possible effects of chromosomal abnormalities on Kisspeptin expression in the study participants.

## Conclusion

The plasma Kisspeptin levels, KISS1 and KISS1R gene expressions in FW and PIW are comparable and do not hold potential for the molecular diagnosis of PFI. However, early menopause is a potential driver for primary infertility through decrease in plasma Kisspeptin and increase in KISS1 gene expression in the blood. This finding justifies the Kisspeptin or KISS1R agonist supplementation in women already showing promise ^[35,67–73]^, which could also be helpful for PIW with history of early menarche. KISS1 gene expression in the blood is central to the role of the Kisspeptin system and may be a significant regulator of its effect within the human blood; it could be used to predict KISS1R gene expression in FW but plasma Kisspeptin levels in PIW.

This study recommends further studies that will unbundle the primary infertility cohort focusing on women with early menarche. To model the multifactorial nature of primary infertility, a highly powered study should study the genetic expression of the Kisspeptin system genes and plasma Kisspeptin levels in multiple reproductive disorders, including women from various clinical settings, to cover for genetic variety. More importantly, future studies should include karyotyping of participants to exclude chromosomal abnormalities that could affect the gene expression levels. However, more outcomes in human studies on KISS1/KISS1R gene expression in other organs like the gonads, pituitary, uterus, and placenta will be very vital in giving holistic evidence on the peripheral actions of Kisspeptins and the role of Kisspeptin in female reproduction. The elucidation of this scientific query will help reveal the whole set of pathophysiologic, diagnostic, and therapeutic implications of Kisspeptin/KISS1/KISS1R in female reproduction.

## Acknowledgements

The authors acknowledge all the staff of the Gynaecology and Family Planning Units of the Department of Obstetrics and Gynaecology, University College Hospital, Ibadan for their assistance with patient education, identification, and recruitment.

## Data Availability and sharing statement

All data used for this study are available upon reasonable request to the authors

## References

1. Venkatesh T, Suresh PS, Tsutsumi R. New insights into the genetic basis of infertility. Application of Clinical Genetics 2014;7:235–43.

2. Mascarenhas MN, Flaxman SR, Boerma T, Vanderpoel S, Stevens GA. National, Regional, and Global Trends in Infertility Prevalence Since 1990: A Systematic Analysis of 277 Health Surveys. PLoS Medicine 2012;9(12):1–12.

3. Abdalla EM, El-Kharadly RN. Pericentric Inversion of Chromosome 9 in a Consanguineous Couple with Molar Pregnancies and Spontaneous Abortions. Laboratory Medicine 2012;43(5):212–6.

4. Elussein EA, Magid YM, Omer MM, Adam I. Clinical patterns and major causes of infertility among Sudanese couples. Trop Doct 2008;38(4):243–4.

5. Punab M, Poolamets O, Paju P, Vihljajev V, Pomm K, Ladva R, et al. Causes of male infertility: a 9-year prospective monocentre study on 1737 patients with reduced total sperm counts. Hum Reprod 2016;humrep;dew284v1.

6. Zegers-Hochschild F, Adamson GD, Dyer S, Racowsky C, de Mouzon J, Sokol R, et al. The International Glossary on Infertility and Fertility Care, 2017†‡§. Human Reproduction 2017;32(9):1786–801.

7. Unuane D, Tournaye H, Velkeniers B, Poppe K. Endocrine disorders & female infertility. Best Practice & Research Clinical Endocrinology & Metabolism 2011;25(6):861–73.

8. Foresta C, Ferlin A, Gianaroli L, Dallapiccola B. Guidelines for the appropriate use of genetic tests in infertile couples. European Journal of Human Genetics 2002;10(5):303–12.

9. Gajbhiye R, Fung JN, Montgomery GW. Complex genetics of female fertility. npj Genomic Medicine [Internet] 2018;3(1). Available from: 10.1038/s41525-018-0068-1

10. Yatsenko SA, Rajkovic A. Genetics of human female infertility†. Biol Reprod 2019;101(3):549–66.

11. Mallepaly R, Butler PR, Herati AS, Lamb DJ. Genetic Basis of Male and Female Infertility. Monographs in Human Genetics 2017;21:1–16.

12. Ayesha B, Ansari A, Lohiya N. An Overview on the Genetic Determinants of Infertility. BJSTR 2018;10(4):7960–4.

13. Zorrilla M, Yatsenko AN. The Genetics of Infertility: Current Status of the Field. Current Genetic Medicine Reports 2013;1(4):247–60.

14. Gottsch ML, Cunningham MJ, Smith JT, Popa SM, Acohido B V., Crowley WF, et al. A role for kisspeptins in the regulation of gonadotropin secretion in the mouse. Endocrinology 2004;145(9):4073–7.

15. Pukos N, Yoseph R, M. McTigue D. To Be or Not to Be: Environmental Factors that Drive Myelin Formation during Development and after CNS Trauma. Neuroglia 2018;1(1):63–90.

16. Greives TJ, Mason AO, Scotti MAL, Levine J, Ketterson ED, Kriegsfeld LJ, et al. Environmental control of kisspeptin: Implications for seasonal reproduction. Endocrinology 2007;148(3):1158–66.

17. Clements MK, McDonald TP, Wang R, Xie G, O’Dowd BF, George SR, et al. FMRFamide-related neuropeptides are agonists of the orphan G-protein-coupled receptor GPR54. Biochemical and Biophysical Research Communications 2001;284(5):1189–93.

18. De Roux N, Genin E, Carel JC, Matsuda F, Chaussain JL, Milgrom E. Hypogonadotropic hypogonadism due to loss of function of the KiSS1-derived peptide receptor GPR54. Proceedings of the National Academy of Sciences of the United States of America 2003;100(19):10972–6.

19. De Tassigny XDA, Fagg LA, Dixon JPC, Day K, Leitch HG, Hendrick AG, et al. Hypogonadotropic hypogonadism in mice lacking a functional Kiss1 gene. Proceedings of the National Academy of Sciences of the United States of America 2007;104(25):10714–9.

20. Gene M suppressor, Lee J hyung, Miele ME, Hicks DJ, Karen K, Trent J, et al. KiSS-1, a Novel Human Malignant Melanoma. Journal of the National Cancer Institute 1996;88(23):1731–7.

21. Luan X, Zhou Y, Wang W, Yu H, Li P, Gan X, et al. Association study of the polymorphisms in the KISS1 gene with central precocious puberty in Chinese girls. European Journal of Endocrinology 2007;157(1):113–8.

22. Lashen H. Investigations for infertility. Current Obstetrics and Gynaecology 2001;11(4):239–44.

23. Edelstam G, Sjösten A, Bjuresten K, Ek I, Wånggren K, Spira J. A new rapid and effective method for treatment of unexplained infertility. Human reproduction (Oxford, England) 2008;23(4):852–6.

24. Kotani M, Detheux M, Vandenbogaerde A, Communi D, Vanderwinden JM, Le Poul E, et al. The Metastasis Suppressor Gene KiSS-1 Encodes Kisspeptins, the Natural Ligands of the Orphan G Protein-coupled Receptor GPR54. Journal of Biological Chemistry 2001;276(37):34631–6.

25. Kuohung W, Kaiser UB. GPR54 and KiSS-1: Role in the regulation of puberty and reproduction. Reviews in Endocrine and Metabolic Disorders 2006;7(4):257–63.

26. Branavan U, Muneeswaran K, Wijesundera WSS, Senanayake A, Chandrasekharan NV, Wijeyaratne CN. Association of Kiss1 and GPR54 Gene Polymorphisms with Polycystic Ovary Syndrome among Sri Lankan Women. BioMed Research International 2019;2019(1996).

27. Okafor IA, Saanu OO, Olayemi O, Omigbodun AO. Characterization of primary female infertility in a Nigerian tertiary hospital: A case-control study. Afr J Reprod Health 2022;26(8):66–82.

28. Glueck CJ, Morrison JA, Wang P, Woo JG. Early and late menarche are associated with oligomenorrhea and predict metabolic syndrome 26years later. Metabolism 2013;62(11):1597–606.

29. Charan J, Biswas T. How to calculate sample size for different study designs in medical research? Indian Journal of Psychological Medicine 2013;35(2):121–6.

30. Vazquez-Alaniz F, Galaviz-Hernandez C, Marchat LA, Salas-Pacheco JM, Chairez-Hernandez I, Guijarro-Bustillos JJ, et al. Comparative expression profiles for KiSS-1 and REN genes in preeclamptic and healthy placental tissues. European Journal of Obstetrics & Gynecology and Reproductive Biology 2011;159(1):67–71.

31. World Health Organization. WHO guidelines on drawing blood: best practices in phlebotomy [Internet]. 2010 [cited 2022 Apr 14];Available from: https://apps.who.int/iris/handle/10665/44294

32. Branavan U, Muneeswaran K, Wijesundera WSS, Senanayake A, Chandrasekharan NV, Wijeyaratne CN. Association of Kiss1 and GPR54 Gene Polymorphisms with Polycystic Ovary Syndrome among Sri Lankan Women. BioMed Research International 2019;2019.

33. de Assis Rodrigues NP, Laganà AS, Zaia V, Vitagliano A, Barbosa CP, de Oliveira R, et al. The role of Kisspeptin levels in polycystic ovary syndrome: a systematic review and meta-analysis. Arch Gynecol Obstet 2019;300(5):1423–34.

34. Emekci Ozay O, Ozay AC, Acar B, Cagliyan E, Seçil M, Küme T. Role of kisspeptin in polycystic ovary syndrome (PCOS). Gynecological Endocrinology 2016;32(9):718–22.

35. Romero-Ruiz A, Skorupskaite K, Gaytan F, Torres E, Perdices-Lopez C, Mannaerts BM, et al. Kisspeptin treatment induces gonadotropic responses and rescues ovulation in a subset of preclinical models and women with polycystic ovary syndrome. Human Reproduction 2019;34(12):2495–512.

36. Varikasuvu SR, Prasad VS, Vamshika V, Satyanarayana M, Panga JR. Circulatory metastin/kisspeptin-1 in polycystic ovary syndrome: a systematic review and meta-analysis with diagnostic test accuracy. Reproductive BioMedicine Online 2019;39(4):685–97.

37. Zeydabadi Nejad S, Ramezani Tehrani F, Zadeh-Vakili A. The Role of Kisspeptin in Female Reproduction. Int J Endocrinol Metab 2017;15(3):e44337.

38. Tang R, Ding X, Zhu J. Kisspeptin and Polycystic Ovary Syndrome. Front Endocrinol 2019;10:298.

39. Liu J, Qu T, Li Z, Yu L, Zhang S, Yuan D, et al. Serum kisspeptin levels in polycystic ovary syndrome: A meta-analysis. J Obstet Gynaecol Res 2021;47(6):2157–65.

40. Blasco V, Pinto FM, Fernández-Atucha A, González-Ravina C, Fernández-Sánchez M, Candenas L. Female infertility is associated with an altered expression of the neurokinin B/neurokinin B receptor and kisspeptin/kisspeptin receptor systems in ovarian granulosa and cumulus cells. Fertility and Sterility 2020;114(4):869–78.

41. Colak E, Ozcimen EE, Erinanç OH, Tohma YA, Ceran MU. Is placental KISS-1 expression associated with first trimester abortion spontaneous? Obstet Gynecol Sci 2020;63(4):490–6.

42. Mayer C, Acosta-Martinez M, Dubois SL, Wolfe A, Radovick S, Boehm U, et al. Timing and completion of puberty in female mice depend on estrogen receptor α-signaling in kisspeptin neurons. Proc Natl Acad Sci USA 2010;107(52):22693–8.

43. Park DW, Lee SK, Hong SR, Han AR, Kwak-Kim J, Yang KM. Expression of Kisspeptin and its Receptor GPR54 in the First Trimester Trophoblast of Women with Recurrent Pregnancy Loss: KISSPEPTIN, GPR54, AND NK CELL IN MISCARRIAGES. American Journal of Reproductive Immunology 2012;67(2):132–9.

44. Pinilla L, Aguilar E, Dieguez C, Millar RP, Tena-Sempere M. Kisspeptins and Reproduction: Physiological Roles and Regulatory Mechanisms. Physiological Reviews 2012;92(3):1235–316.

45. Reynolds RM, Logie JJ, Roseweir AK, McKnight AJ, Millar RP. A role for kisspeptins in pregnancy: facts and speculations. Reproduction 2009;138(1):1–7.

46. Wu S, Zhang H, Tian J, Liu L, Dong Y, Mao T. Expression of kisspeptin/GPR54 and PIBF/PR in the first trimester trophoblast and decidua of women with recurrent spontaneous abortion. Pathology - Research and Practice 2014;210(1):47–54.

47. Heung MMS, Jin S, Tsui NBY, Ding C, Leung TY, Lau TK, et al. Placenta-Derived Fetal Specific mRNA Is More Readily Detectable in Maternal Plasma than in Whole Blood. PLoS ONE 2009;4(6):e5858.

48. Makri A, Msaouel P, Petraki C, Milingos D, Protopapas A, Liapi A, et al. KISS1/KISS1R Expression in Eutopic and Ectopic Endometrium of Women Suffering from Endometriosis. In Vivo 2012;26(1):119–27.

49. Abebe MS, Afework M, Abaynew Y. Primary and secondary infertility in Africa: systematic review with meta-analysis. Fertil Res and Pract 2020;6(1):20.

50. Akalewold M, Yohannes GW, Abdo ZA, Hailu Y, Negesse A. Magnitude of infertility and associated factors among women attending selected public hospitals in Addis Ababa, Ethiopia: a cross-sectional study. BMC Women’s Health 2022;22(1):11.

51. Okafor IA, Saanu OO, Olayemi O, Omigbodun A. The role of Kisspeptin, KISS1 and KISS1R genes in primary female infertility: a case-control study of a Nigerian population. The FASEB Journal 2022;36(S1):fasebj.2022.36.S1.R4495.

52. Rafique N, Latif R. Serum kisspeptin levels in normal and overweight Saudi females and its relation with anthropometric indices. Annals of Saudi Medicine 2015;35(2):157–60.

53. Rashad NM, Al-sayed RM, Yousef MS, Saraya YS. Kisspeptin and body weight homeostasis in relation to phenotypic features of polycystic ovary syndrome; metabolic regulation of reproduction. Diabetes & Metabolic Syndrome: Clinical Research & Reviews 2019;13(3):2086–92.

54. Tolson KP, Garcia C, Yen S, Simonds S, Stefanidis A, Lawrence A, et al. Impaired kisspeptin signaling decreases metabolism and promotes glucose intolerance and obesity. J Clin Invest 2014;124(7):3075–9.

55. Holmes D. Kisspeptin signalling linked to obesity. Nat Rev Endocrinol 2014;10(9):511–511.

56. Topaloglu AK, Turan I. Genetic Etiology of Idiopathic Hypogonadotropic Hypogonadism. Endocrines 2021;3(1):1–15.

57. Abbara A, Dhillo WS. Makorin rings the kisspeptin bell to signal pubertal initiation. Journal of Clinical Investigation 2020;130(8):3957–60.

58. Chen CY, Chou YY, Wu YM, Lin CC, Lin SJ, Lee CC. Phthalates may promote female puberty by increasing kisspeptin activity. Human Reproduction 2013;28(10):2765–73.

59. Cintra RG, Wajnsztejn R, Trevisan CM, Zaia V, Laganà AS, Bianco B, et al. Kisspeptin Levels in Girls with Precocious Puberty: A Systematic Review and Meta-Analysis. Horm Res Paediatr 2020;93(11–12):589–98.

60. Roa J, Barroso A, Ruiz-Pino F, Vázquez MJ, Seoane-Collazo P, Martínez-Sanchez N, et al. Metabolic regulation of female puberty via hypothalamic AMPK–kisspeptin signaling. Proc Natl Acad Sci USA [Internet] 2018 [cited 2022 May 12];115(45). Available from: https://pnas.org/doi/full/10.1073/pnas.1802053115

61. De Roux N, Genin E, Carel JC, Matsuda F, Chaussain JL, Milgrom E. Hypogonadotropic hypogonadism due to loss of function of the KiSS1-derived peptide receptor GPR54. Proceedings of the National Academy of Sciences of the United States of America 2003;100(19):10972–6.

62. Funes S, Hedrick JA, Vassileva G, Markowitz L, Abbondanzo S, Golovko A, et al. The KiSS-1 receptor GPR54 is essential for the development of the murine reproductive system. Biochemical and Biophysical Research Communications 2003;312(4):1357–63.

63. Seminara SB, Messager S, Chatzidaki EE, Thresher RR, Acierno JJ, Shagoury JK. The GPR54 gene as a regulator of puberty. N Engl J Med 2003;349(17):1614–27.

64. Silveira LG, Noel SD, Silveira-Neto AP, Abreu AP, Brito VN, Santos MG, et al. Mutations of the KISS1 gene in disorders of puberty. Journal of Clinical Endocrinology and Metabolism 2010;95(5):2276–80.

65. Teles MG, Silveira LFG, Tusset C, Latronico AC. New genetic factors implicated in human GnRH-dependent precocious puberty: The role of kisspeptin system. Molecular and Cellular Endocrinology 2011;346(1–2):84–90.

66. Towne B, Czerwinski SA, Demerath EW, Blangero J, Roche AF, Siervogel RM. Heritability of age at menarche in girls from the Fels Longitudinal Study. Am J Phys Anthropol 2005;128(1):210–9.

67. Dhillo WS, Chaudhri OB, Thompson EL, Murphy KG, Patterson M, Ramachandran R, et al. Kisspeptin-54 Stimulates Gonadotropin Release Most Potently during the Preovulatory Phase of the Menstrual Cycle in Women. The Journal of Clinical Endocrinology & Metabolism 2007;92(10):3958–66.

68. George JT, Seminara SB. Kisspeptin and the Hypothalamic Control of Reproduction: Lessons from the Human. Endocrinology 2012;153(11):5130–6.

69. Jayasena CN, Nijher GMK, Abbara A, Murphy KG, Lim A, Patel D, et al. Twice-Weekly Administration of Kisspeptin-54 for 8 Weeks Stimulates Release of Reproductive Hormones in Women With Hypothalamic Amenorrhea. Clin Pharmacol Ther 2010;88(6):840–7.

70. Abbara A, Jayasena CN, Christopoulos G, Narayanaswamy S, Izzi-Engbeaya C, Nijher GMK, et al. Efficacy of Kisspeptin-54 to Trigger Oocyte Maturation in Women at High Risk of Ovarian Hyperstimulation Syndrome (OHSS) During In Vitro Fertilization (IVF) Therapy. The Journal of Clinical Endocrinology & Metabolism 2015;100(9):3322–31.

71. Abbara A, Clarke S, Islam R, Prague JK, Comninos AN, Narayanaswamy S, et al. A second dose of kisspeptin-54 improves oocyte maturation in women at high risk of ovarian hyperstimulation syndrome: a Phase 2 randomized controlled trial. Human Reproduction 2017;32(9):1915–24.

72. Jayasena CN, Abbara A, Comninos AN, Nijher GMK, Christopoulos G, Narayanaswamy S, et al. Kisspeptin-54 triggers egg maturation in women undergoing in vitro fertilization. J Clin Invest 2014;124(8):3667–77.

73. Qin L, Sitticharoon C, Petyim S, Keadkraichaiwat I, Sririwichitchai R, Maikeaw P, et al. Roles of kisspeptin in IVF/ICSI-treated infertile women and in human granulosa cells. Exp Biol Med (Maywood) 2021;246(8):996–1010.

74. Calley JL, Dhillo WS. Effects of the Hormone Kisspeptin on Reproductive Hormone Release in Humans. 2014.

75. Ruohonen ST, Poutanen M, Tena-Sempere M. Role of kisspeptins in the control of the hypothalamic-pituitary-ovarian axis: old dogmas and new challenges. Fertility and Sterility 2020;114(3):465–74.

76. Adachi S, Yamada S, Takatsu Y, Matsui H, Kinoshita M, Takase K, et al. Involvement of Anteroventral Periventricular Metastin/Kisspeptin Neurons in Estrogen Positive Feedback Action on Luteinizing Hormone Release in Female Rats. J Reprod Dev 2007;53(2):367–78.

77. Clarkson J, Herbison AE. Postnatal Development of Kisspeptin Neurons in Mouse Hypothalamus; Sexual Dimorphism and Projections to Gonadotropin-Releasing Hormone Neurons. Endocrinology 2006;147(12):5817–25.

78. Mattam U, Talari N, Thiriveedi V, Fareed M, Velmurugan S, Mahadev K, et al. Aging reduces kisspeptin receptor (GPR54) expression levels in the hypothalamus and extra-hypothalamic brain regions. Exp Ther Med 2021;22(3):1019.

79. Smith JT, Cunningham MJ, Rissman EF, Clifton DK, Steiner RA. Regulation of Kiss1 Gene Expression in the Brain of the Female Mouse. Endocrinology 2005;146(9):3686–92.

80. Smith JT, Dungan HM, Stoll EA, Gottsch ML, Braun RE, Eacker SM, et al. Differential Regulation of KiSS-1 mRNA Expression by Sex Steroids in the Brain of the Male Mouse. Endocrinology 2005;146(7):2976–84.

81. Smith JT, Clay CM, Caraty A, Clarke IJ. KiSS-1 Messenger Ribonucleic Acid Expression in the Hypothalamus of the Ewe Is Regulated by Sex Steroids and Season. Endocrinology 2007;148(3):1150–7.

82. Shibata M, Friedman RL, Ramaswamy S, Plant TM. Evidence That Down Regulation of Hypothalamic KiSS-1 Expression is Involved in the Negative Feedback Action of Testosterone to Regulate Luteinising Hormone Secretion in the Adult Male Rhesus Monkey (Macaca mulatta). J Neuroendocrinol 2007;19(6):432–8.

83. Yamada S, Uenoyama Y, Kinoshita M, Iwata K, Takase K, Matsui H, et al. Inhibition of Metastin (Kisspeptin-54)-GPR54 Signaling in the Arcuate Nucleus-Median Eminence Region during Lactation in Rats. Endocrinology 2007;148(5):2226–32.

84. Sullivan-Pyke C, Haisenleder DJ, Senapati S, Nicolais O, Eisenberg E, Sammel MD, et al. Kisspeptin as a new serum biomarker to discriminate miscarriage from viable intrauterine pregnancy. Fertility and Sterility 2018;109(1):137–141.e2.

85. Gorbunova O, Shirshev S. The effect of kisspeptin on the functional activity of peripheral blood monocytes and neutrophils in the context of physiological pregnancy. Journal of Reproductive Immunology 2022;151:103621.

86. Shahab M, Lippincott M, Chan YM, Davies A, Merino PM, Plummer L, et al. Discordance in the Dependence on Kisspeptin Signaling in Mini Puberty vs Adolescent Puberty: Human Genetic Evidence. The Journal of Clinical Endocrinology & Metabolism 2018;103(4):1273–6.

